# Mixed ancestry analysis of whole-genome sequencing identifies *TBX5* and *PTK7* as susceptibility genes for posterior urethral valves

**DOI:** 10.1101/2021.08.09.21261801

**Authors:** Melanie MY Chan, Omid Sadeghi-Alavijeh, Filipa M Lopes, Alina C Hilger, Horia C Stanescu, Catalin D Voinescu, Glenda M Beaman, William G Newman, Marcin Zaniew, Stefanie Weber, John O Connolly, Dan Wood, Alexander Stuckey, Athanasios Kousathanas, Genomics England Research Consortium, Robert Kleta, Adrian S Woolf, Detlef Bockenhauer, Adam P Levine, Daniel P Gale

**Affiliations:** Department of Renal Medicine, University College London, London, NW3 2PF, UK.; Division of Cell Matrix Biology & Regenerative Medicine, School of Biological Sciences, Faculty of Biology, Medicine and Health, University of Manchester, Manchester, M13 9WL, UK.; Royal Manchester Children’s Hospital, Manchester University NHS Foundation Trust, Manchester Academic Health Science Centre, Manchester, M13 9WL, UK.; Children’s Hospital, University of Bonn, 53113 Bonn, Germany.; Institute of Human Genetics, University of Bonn, 53127 Bonn, Germany.; Manchester Centre for Genomic Medicine, Manchester University NHS Foundation Trust, Manchester, M13 9WL, UK.; Evolution and Genomic Sciences, School of Biological Sciences, University of Manchester, Manchester, M13 9PL, UK.; Department of Pediatrics, University of Zielona Góra, 56-417 Zielona Góra, Poland.; Department of Pediatric Nephrology, University of Marburg, 35037 Marburg, Germany.; Department of Adolescent Urology, University College London Hospitals NHS Foundation Trust, London, NW1 2BU, UK.; Genomics England, Queen Mary University of London, London, EC1M 6BQ, UK.; Nephrology Department, Great Ormond Street Hospital for Children NHS Foundation Trust, London, WC1N 3JH, UK.; Research Department of Pathology, University College London, London, WC1E6DD, UK.

**Keywords:** whole-genome sequencing, posterior urethral valves, PUV, GWAS, genetics, mixed-ancestry, CAKUT

## Abstract

Posterior urethral valves (PUV) are the commonest cause of end-stage renal disease in children, but the genetic architecture of this rare disorder remains largely unknown. We analyzed whole-genome sequencing (WGS) data from 132 unrelated PUV cases and 23,727 controls of mixed ancestry and identified statistically significant associations with common variants at 12q24.21 (*P*=7.8x10^-12^; OR 0.4) and rare variants at 6p21.1 (*P*=2x10^-8^; OR 7.2), that were replicated in an independent European cohort. Bayesian fine mapping and functional annotation mapped these loci to the transcription factor *TBX5* and planar cell polarity gene *PTK7*, respectively, with the encoded proteins detected in the normal human developing urinary tract. These findings represent the first known genetic associations of PUV, providing novel insights into the underlying biology of this poorly understood disorder and demonstrate that a mixed ancestry WGS approach can increase power for disease locus discovery and facilitate fine-mapping of causal variants.

## Introduction

Posterior urethral valves (PUV) are the commonest cause of end-stage renal disease (ESRD) in children, affecting 1 in 4,000 male births^1, 2^ and resulting in congenital bladder outflow obstruction. PUV is a uniquely male disorder, with a quarter of those affected developing ESRD (i.e., requirement for dialysis or kidney transplantation) before the age of 30 years.^3, 4^ PUV is often associated with renal dysplasia, vesicoureteral reflux (VUR) and bladder dysfunction which are poor prognostic factors for renal survival.^3^ Management involves endoscopic valve ablation in infancy to relieve the obstruction, however the majority of affected children have long-term sequelae related to ongoing bladder dysfunction.^5^

During embryogenesis, the bladder, prostate, and urethra develop from the endoderm-derived urogenital sinus, while the distal mesonephric (Wolffian) duct forms the base of the bladder (trigone) before integrating into the prostatic urethra to become the ejaculatory ducts in males.^6^ Abnormal integration of the mesonephric duct into the posterior urethra or persistence of the urogenital membrane have both been proposed as possible mechanisms underlying PUV,^6^ but the exact biological processes involved remain poorly understood.

Although usually sporadic, familial clustering and twin studies suggest a genetic component underlying PUV.^7–10^ Monogenic causes of anatomical and functional congenital bladder outflow obstruction have been described (*BNC2* in urethral stenosis and atypical PUV,^11^ *HPSE2* and *LRIG2* in urofacial syndrome,^12, 13^ *CHRM3* in prune-belly like syndrome,^14^ and *MYOCD* in congenital megabladder^15^), however a monogenic etiology for classical PUV has not been identified. Case reports and microarray studies have linked PUV with chromosomal abnormalities^16–18^ and rare copy number variants (CNVs),^19–22^ suggesting structural variation may be important, but the underlying genetic architecture has so far remained largely uncharacterized.

Here, we use whole-genome sequencing (WGS) in a large mixed ancestry cohort to investigate how common, low-frequency, and rare single-nucleotide and structural variation contribute to this complex disorder. Through genome-wide association analysis we identify two novel genetic loci that implicate *TBX5* (T-Box Transcription Factor 5) and *PTK7* (Protein Tyrosine Kinase 7) and show that the encoded proteins are detected in the normal human developing urinary tract. In addition, we demonstrate that a well-controlled diverse ancestry WGS approach can increase power for disease locus discovery and facilitate the fine-mapping of causal variants.

## Results

We analyzed WGS data from 132 unrelated male probands with PUV and 23,727 non-PUV controls (unaffected relatives without known kidney disease), recruited to the UK 100,000 Genomes Project (100KGP)^23^ (see Fig. S1 for study workflow). The available dataset (version 10) combined WGS data, clinical phenotypes standardized using Human Phenotype Ontology (HPO) codes, and comprehensive hospital clinical records for 89,139 individuals with cancer, rare disease, and their unaffected relatives. None of the cases included had received a definitive genetic diagnosis through the clinical arm of the 100KGP. Two individuals had a pathogenic and likely pathogenic variant affecting *HNF1B* and *FOXC1,* respectively, but these were not considered causal for PUV (see Supplemental Note). Given the small number of recruited cases with this rare disorder, we chose to jointly analyze individuals from diverse ancestral backgrounds, thereby preserving sample size and boosting power. To mitigate confounding due to population structure whilst using this mixed ancestry approach we employed two strategies. First, we carried out ancestry-matching of cases and controls using weighted principal components (Fig. S2), and second, we utilized a generalized logistic mixed model to account for relatedness between individuals. Clinical characteristics and genetic ancestry of the cases and controls are detailed in Table S1.

**Table 1.** Association statistics for significant genome-wide loci. The lead variant with the lowest *P* value at each locus is shown with genome-wide significance defined as *P*<5×10^-8^. Genomic positions are with reference to GRCh38. Discovery *P* values were derived using SAIGE generalized logistic mixed model association test and replication *P* values using a one-sided Cochran Armitage Trend Test. CHR, chromosome; POS, position; OR, odds ratio; CI, confidence interval; EAF, effect allele frequency.

| Lead variant | CHR:POS | Effect Allele | Closest gene | $P$ value | | OR (95% CI) | | Case EAF | | Control EAF | |
| --- | --- | --- | --- | --- | --- | --- | --- | --- | --- | --- | --- |
|  |  |  |  | Discovery | Replication | Discovery | Replication | Discovery | Replication | Discovery | Replication |
| rs10774740 | chr12:114228397 | T | *TBX5* | $7.81 \times 10^{-12}$ | $1.9 \times 10^{-3}$ | 0.40<br>(0.31-0.52) | 0.78<br>(0.67-0.91) | 0.19 | 0.31 | 0.37 | 0.36 |
| rs144171242 | chr6:43120356 | G | *PTK7* | $2.02 \times 10^{-8}$ | $4.5 \times 10^{-3}$ | 7.20<br>(4.08-12.70) | 2.17<br>(1.25-3.76) | 0.05 | 0.02 | 0.01 | 0.01 |

### Variation at 12q24.21 and 6p21.1 is associated with PUV

To determine the contribution of common and low-frequency variation to PUV, we carried out a genome-wide association analysis of 17,091,503 variants with MAF > 0.1%. The genomic inflation factor (λ) of 1.05 confirmed population stratification was well controlled in this mixed ancestry cohort (Fig. S3). Statistically significant (*P*<5×10^-8^) associations were detected at two loci (Fig. 2 and Table 1). At 12q24.21, the lead intergenic variant (rs10774740) was common (MAF 0.37) and reached *P*=7.81×10^-12^ (OR 0.40; 95% CI 0.31-0.52; Fig. 3A). A rare (MAF 0.007) variant (rs144171242) at 6p21.1, located in an intron of *PTK7,* was also significant at *P*=2.02×10^-8^ (OR 7.20; 95% CI 4.08-12.70; Fig. 4A). Table S6 details the summary statistics for variants with *P*<10^-5^. Conditional analysis did not identify secondary independent signals at either locus and epistasis was not detected between the two lead variants (*P*=0.10). Gene and gene-set analysis was carried out to assess the joint effect of common and low-frequency variants and identify potential functional pathways associated with PUV, however, no genes (Table S7) or pathways (Table S8) reached statistical significance after correction for multiple testing.

**Figure 1.**
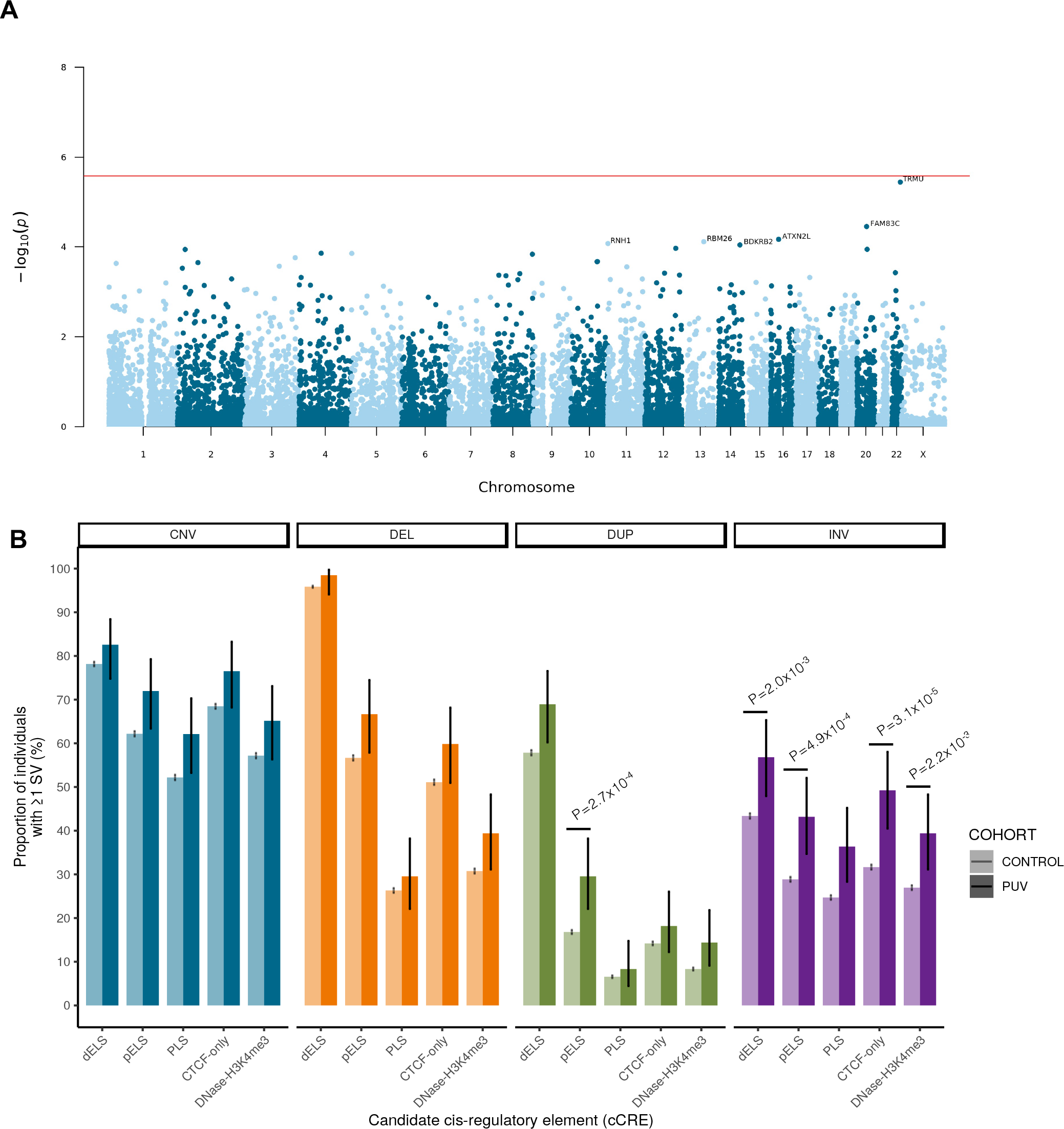
Rare variant analysis. **A**, Manhattan plot of exome-wide rare coding variant analysis demonstrating no significant gene-based enrichment after correction for multiple testing. Chromosomal position (GRCh38) is shown on the x axis and strength of association using a –log_10_(P) scale on the y axis. Each dot represents a gene. The red line indicates the Bonferroni adjusted threshold for exome-wide significance (*P* = 2.58×10^-6^). Genes with *P* < 10^-4^ are labelled. **B**, The proportion of individuals with ≥1 rare autosomal structural variant intersecting with an ENCODE cCRE in cases and controls was enumerated using a two-sided Fisher’s exact test. Note that inversions affecting cCRE are enriched in PUV. Vertical black bars indicate 95% confidence intervals. Unadjusted *P* values shown are significant after correction for multiple testing (*P* < 2.5×10^-3^). CNV, copy number variant; DEL, deletion; DUP, duplication; INV, inversion; PUV, posterior urethral valves; dELS, distal enhancer-like signature; pELS, proximal enhancer-like signature; PLS, promoter-like signature; cCRE, candidate cis-regulatory element.

**Figure 2.**
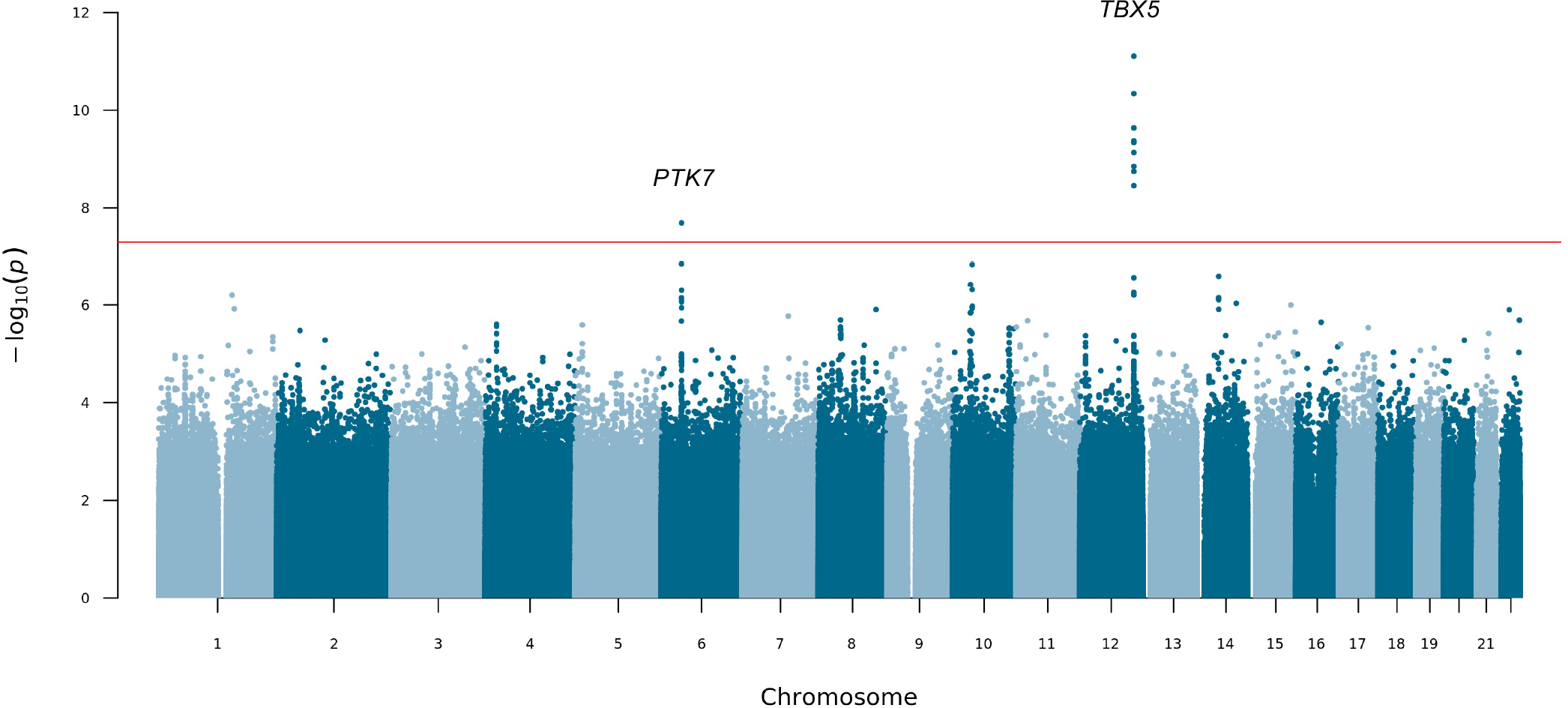
Manhattan plot for mixed-ancestry GWAS. A genome-wide single-variant association study was carried out in 132 unrelated PUV cases and 23,727 controls for 17,091,503 variants with MAF > 0.1%. Chromosomal position (GRCh38) is denoted along the x axis and strength of association using a –log_10_(*P*) scale on the y axis. Each dot represents a variant. The red line indicates the Bonferroni adjusted threshold for genome-wide significance (*P* < 5×10^-8^). The gene in closest proximity to the lead variant at significant loci are listed.

### 12q24.21 and 6p21.1 replicate in an independent cohort

We next carried out a replication study in an independent European cohort consisting of 398 individuals with PUV: 336 from Poland and Germany, partially recruited through the CaRE for LUTO (Cause and Risk Evaluation for Lower Urinary Tract Obstruction) Study, and 62 from the UK. 10,804 European individuals recruited to the cancer arm of the 100KGP were used as controls. The UK PUV patients and the 100KGP cancer control cohort had not been included in the discovery analyses. The lead variants at the top four loci with *P*<5×10^-7^ were tested for replication. Association at both genome-wide significant lead variants was replicated although with smaller effect sizes (Table 1): rs10774740 (*P*=1.9×10^-3^; OR 0.78; 95% CI 0.67-0.91) and rs144171242 (*P*=4.5×10^-3^; OR 2.17; 95% CI 1.25-3.76). Two further loci with suggestive evidence of association (10q11.2; rs1471950716; *P*=1.45×10^-7^ and 14q21.1; rs199975325; *P*=2.52×10^-7^) did not replicate (Table S9).

### Mixed ancestry analysis increases power for discovery

To ascertain whether the observed associations were being driven by a specific ancestry group, we next repeated the GWAS using a subgroup of genetically defined European individuals (88 cases and 17,993 controls) and 15,447,192 variants with MAF > 0.1%. The 12q24.21 locus remained genome-wide significant (Fig. S4), however the lead variant (rs2555009) in the region showed weaker association (*P*=4.02×10^-8^; OR 0.43; 95% CI 0.12-0.73) than rs10774740, the lead variant in the mixed ancestry analysis (Table 10). Interestingly the two variants were not in strong linkage disequilibrium (LD; r^2^=0.54). The lead variant at 6p21.1 from the mixed ancestry analysis did not reach genome-wide significance in the European-only study (rs144171242; *P*=3.60×10^-5^; OR 5.90; 95% CI 2.88-12.11) suggesting that this signal may be driven partly by non-Europeans. *P* values and effect sizes were strongly correlated between the mixed ancestry and European-only GWAS (Fig. S5) demonstrating that inclusion of mixed ancestry individuals to increase sample size can be an effective way to boost power and discover novel loci, even in a small cohort.

As the numbers of African, South Asian, and admixed ancestry individuals were too small to reliably carry out subgroup association analyses and subsequent meta-analysis, we instead compared ancestry-specific allele frequencies, effect sizes and directions for the two lead variants. Interestingly, rs10774740 (T) had a higher allele frequency in individuals of African ancestry (MAF 0.74) compared to European (MAF 0.37) and South Asian (MAF 0.35) populations, however the effect size and direction was similar between the groups (Fig. S6). rs144171242 (G) was present at a lower allele frequency in South Asian (MAF 0.002) compared to European (MAF 0.008) individuals and was not seen in the African ancestry group. The effect size of this rare variant was higher in the South Asian than European population (Fig. S6), which may explain why it only reached genome-wide significance after inclusion of South Asian individuals. Finally, comparison with population allele frequencies fro gnomAD^24^ demonstrated that although there is large variation in the allele frequency of rs10774740 between ancestries this is away from, not towards, the case allele frequency and confirms that the detected associations are not being driven by differences in allele frequency between populations (Fig. S6).

### Fine-mapping predicts lead variants to be causal

WGS enables further interrogation of loci of interest at high resolution. We therefore repeated the mixed ancestry analysis at each genome-wide significant locus using all variants with minor allele count ≥ 3, to determine whether additional ultra-rare variants might be driving the observed association signals. Both rs10774740 at 12q24.21 and rs144171242 at 6p21.1 remained most strongly associated, suggesting they are likely to be causal. Comparison of the different LD patterns seen across African, European, and South Asian population groups at these loci demonstrate how a combined ancestry approach can leverage differences in LD to improve the fine mapping of causal variants (Fig. S7).

We next applied the Bayesian fine-mapping tool PAINTOR^25^ which integrates the strength of association, LD patterns and functional annotations to derive the posterior probability of a variant being causal. Using this alternative statistical approach, both lead variants were identified as having high probability of being causal: rs10774740 (posterior probability [PP] with no annotations 0.77, PP with annotations >0.99) and rs144171242 (PP with no annotations 0.83, PP with annotations >0.99). Conservation and ChIP-seq transcription factor binding clusters had the largest impact on posterior probabilities at 12q24.21 and 6p21.1, respectively. Validation of the lead variants using statistical fine mapping illustrates that the increased sensitivity and improved resolution of WGS compared with genotyping arrays may permit the direct identification of underlying causal variants, particularly in the context of examining rarer variants and multiple ancestries for which imputation performance may be limiting.^26, 27^

### Functional annotation implicates *TBX5* and *PTK7*

To explore the functional relevance of these loci we next interrogated publicly available datasets via UCSC Genome Browser^28^ and used Functional Mapping and Annotation (FUMA)^29^ to prioritize candidate genes. Given the urinary tract is derived from both embryonic mesoderm and endoderm, where possible we used experimental data obtained from male H1 BMP4-derived mesendoderm cultured cells.

The common, non-coding, intergenic lead variant (rs10774740) at the 12q24.21 locus is predicted to be deleterious (CADD score 15.54) and intersects with a conserved element (chr12:114228397-114228414; logarithm of odds score 33) that is suggestive of a putative transcription factor binding site (TFBS) (Fig. 3B), however review of experimentally defined TF binding profiles^30^ did not identify any known interactions with DNA-binding motifs at this position. Interrogation of epigenomic data from ENCODE^31^ revealed rs10774740 is located ∼35bp away from a candidate cis-regulatory element (cCRE, EH38E1646218), which although has low-DNase activity in mesendoderm cells, displays a distal enhancer-like signature in cardiac myocytes. We did not identify any *cis*-eQTL associations with rs10774740, but using experimental Hi-C data generated from H1 BMP4-derived mesendoderm cells^32, 33^ we were able to determine that this locus is within the same topologically-associated domain (TAD) as the transcription factor *TBX5* (Fig. 4C). Chromatin interactions mapped this intergenic locus directly to the promoter of *TBX5* (false discovery rate [FDR] 2.80×10^-13^, Fig. 4D).

**Figure 3.**
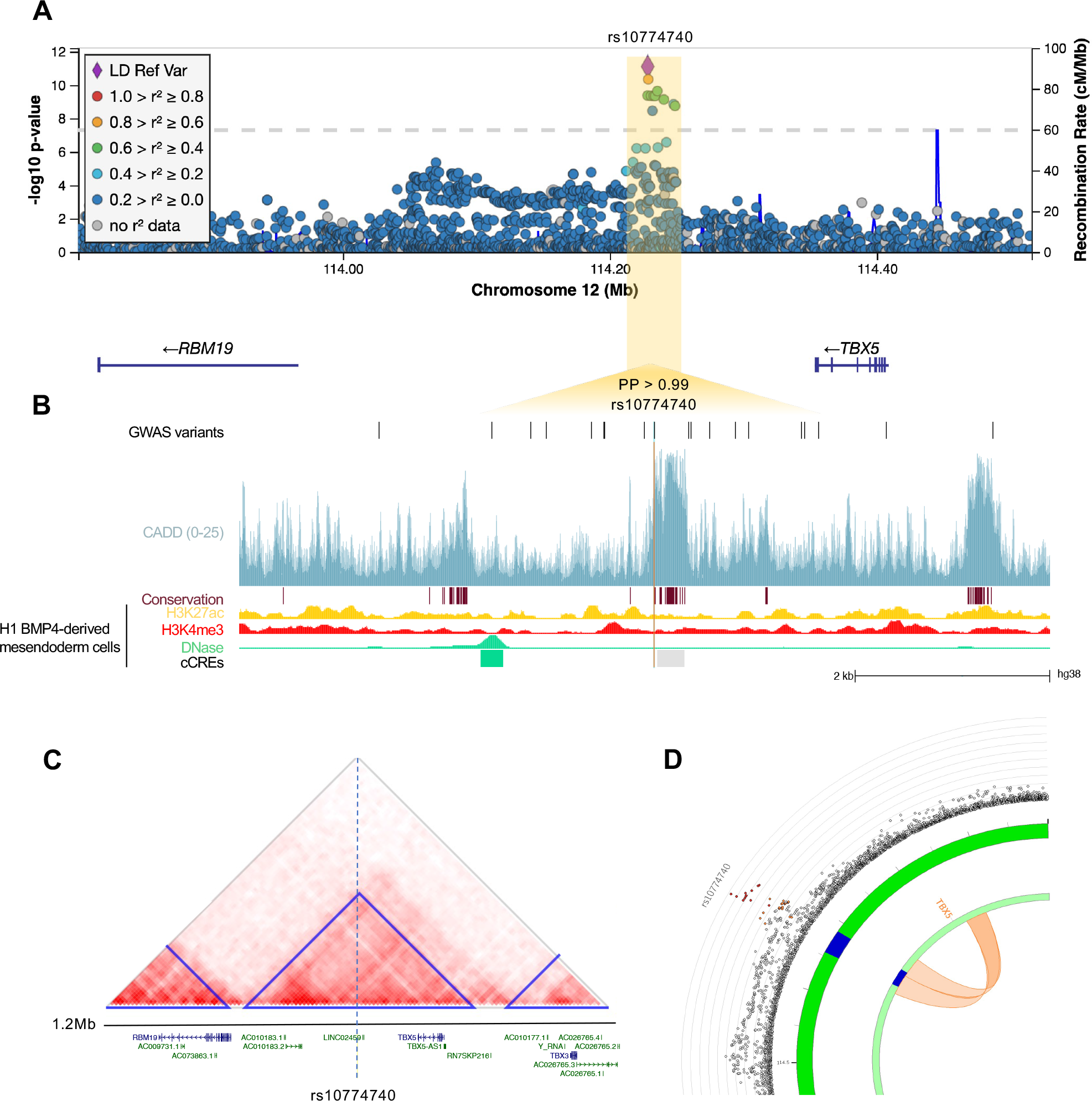
12q24.21. **A**, Regional association plot with chromosomal position (GRCh38) denoted along the x axis and strength of association using a –log_10_(*P*) scale on the y axis. The lead variant (rs10774740) is represented by a purple diamond. Variants are colored based on their linkage disequilibrium (LD) with the lead variant using 1000 Genomes data from all population groups. **B**, Functional annotation of the lead prioritized variant rs10774740 showing intersection with CADD score (v1.6), PhastCons conserved elements from 100 vertebrates, and ENCODE H3K27ac ChIP-seq, H3K4me3 ChIP-seq and DNase-seq from mesendoderm cells. ENCODE cCREs active in mesendoderm are represented by shaded boxes; low-DNase (grey), DNase-only (green). GWAS variants with *P* < 0.05 are shown. Note that rs10774740 has a relatively high CADD score for a non-coding variant and intersects with a highly conserved region. **C**, Heatmap of Hi-C interactions from H1 BMP4-derived mesendoderm cells demonstrating that rs10774740 is located within the same topologically associating domain (TAD) as *TBX5*. TADs are represented by blue triangles. Protein-coding genes are denoted in blue, non-coding genes in green. **D**, Zoomed in circos plot illustrating significant chromatin interactions between 12q24.21 and the promoter of *TBX5*. The outer layer represents a Manhattan plot with variants plotted against strength of association. Only variants with *P* < 0.05 are displayed. Genomic risk loci are highlighted in blue in the second layer. Significant chromatin loops detected in H1 BMP4-derived mesendoderm cultured cells are represented in orange. PP, posterior probability derived using PAINTOR; cCRE, candidate cis-regulatory element.

**Figure 4.**
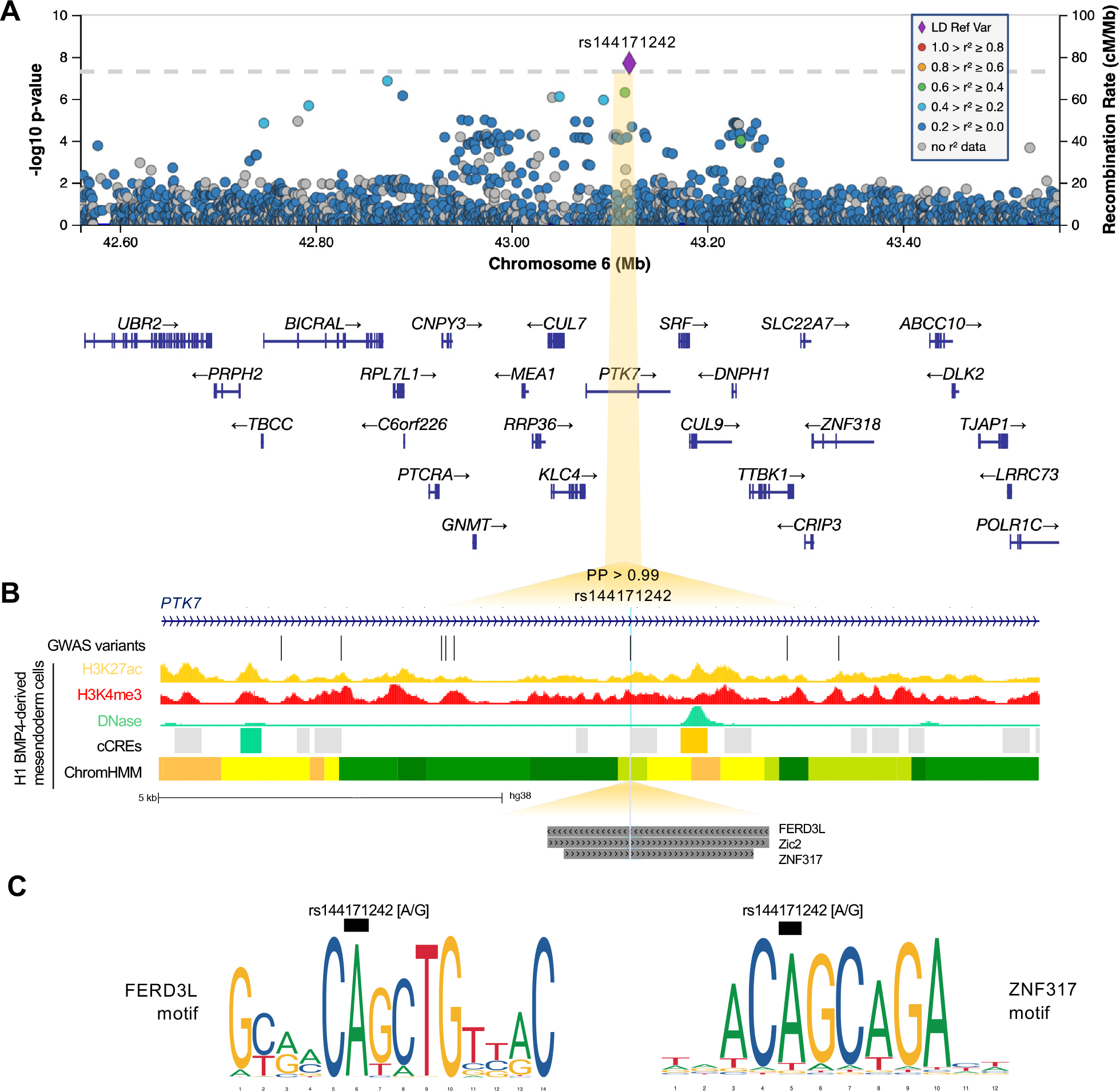
6p21.1. **A**, Regional association plot with chromosomal position (GRCh38) along the x axis and strength of association using a –log_10_(*P*) scale on the y axis. The lead variant (rs144171242) is represented by a purple diamond. Variants are colored based on their linkage disequilibrium (LD) with the lead variant using 1000 Genomes data from all population groups. **B**, Functional annotation of lead prioritized variant rs144171242 showing intersection with ENCODE H3K27ac ChIP-seq, H3K4me3 ChIP-seq and DNase-seq from mesendoderm cells. ENCODE cCREs active in mesendoderm are represented by shaded boxes; low-DNase (grey), DNase-only (green) and distal enhancer-like (orange). ChromHMM illustrates predicted chromatin states using Roadmap Epigenomics imputed 25-state model for mesendoderm cells; active enhancer (orange), weak enhancer (yellow), strong transcription (green), transcribed and weak enhancer (lime green). Predicted transcription factor binding sites (TFBS) from the JASPAR 2020 CORE collection are indicated by dark grey shaded boxes. GWAS variants with *P* < 0.05 are shown. Note that rs144171242 intersects with both a predicted regulatory region and TFBS. **C**, Sequence logos representing the DNA-binding motifs of transcription factors FERD3L and ZNF317. The black boxes indicate where the risk allele [G] may disrupt binding. PP, posterior probability derived using PAINTOR; cCREs, candidate cis-regulatory elements.

At the 6p21.1 locus, the non-coding lead variant (rs144171242) is in an intron of the inactive tyrosine kinase *PTK7*. This rare variant has a low CADD score (0.93) and lacks any known eQTL or relevant chromatin interaction associations. Interrogation of epigenomic annotations from ENCODE^31^ revealed rs144171242 intersects a cCRE (EH38E2468259) with low DNase activity in mesendoderm cells, but with a distal enhancer-like signature in neurons (Fig. 4B). NIH Roadmap Epigenomics Consortium^34^ data suggests rs144171242 may have regulatory activity in mesendoderm cells, classifying this region as transcribed/weak enhancer (12TxEnhW) using the imputed ChromHMM 25-chromatin state model (Fig. 4B). In addition, interrogation of the JASPAR 2020^30^ database of experimentally defined TF binding profiles revealed rs144171242 intersects with the DNA-binding motifs of FERD3L, ZNF317 and Zic2 (Fig. 4C), suggesting rs144171242 may potentially affect *PTK7* expression via disruption of TF binding (Fig. 4D).

### rs10774740 is associated with prostate cancer

Interrogation of the NHGR/EBI GWAS Catalog^35^ revealed the risk allele rs10774740 (G) is associated with prostate cancer aggressiveness^36^ (*P*=3×10^-10^; OR 1.14; 95% CI 1.09-1.18). PheWAS data from the UK Biobank demonstrated the protective allele rs10774740 (T) also has a protective effect in female genitourinary phenotypes: urinary incontinence (*P*=8.3×10^-12^; OR 0.90; 95% CI 0.87-0.92), female stress incontinence (*P*=7.9×10^-10^; OR 0.89; 95% CI 0.85-0.92), genital prolapse (*P*=1.1×10^-9^; OR 0.92; CI 0.89-0.94) and symptoms involving the female genital tract (*P*=1.7×10^-8^; OR 0.90; 95% CI 0.87-0.94). No known GWAS or PheWAS associations were identified for rs144171242.

### TBX5 and PTK7 proteins are detected in the developing urinary tract

To determine whether TBX5 and PTK7 proteins can be detected during urinary tract development, immunohistochemistry was undertaken in a seven-week gestation normal human embryo (Fig. 5A). At this stage of development, the urogenital sinus is a tube comprised of epithelia that will differentiate into urothelial cells of the proximal urethra and the urinary bladder. Uroplakin 1B, a water-proofing protein, was detected in urogenital sinus epithelia (Fig. 5B). PTK7 was detected in epithelia lining the urogenital sinus, and intensely in stromal-like cells surrounding the mesonephric ducts (Fig. 5C). TBX5 was detected in a nuclear pattern in a subset of epithelial cells lining the urogenital sinus (Fig. 5D). Omission of primary antibodies resulted in absent signals, as expected (Fig. 5E).

**Figure 5.**
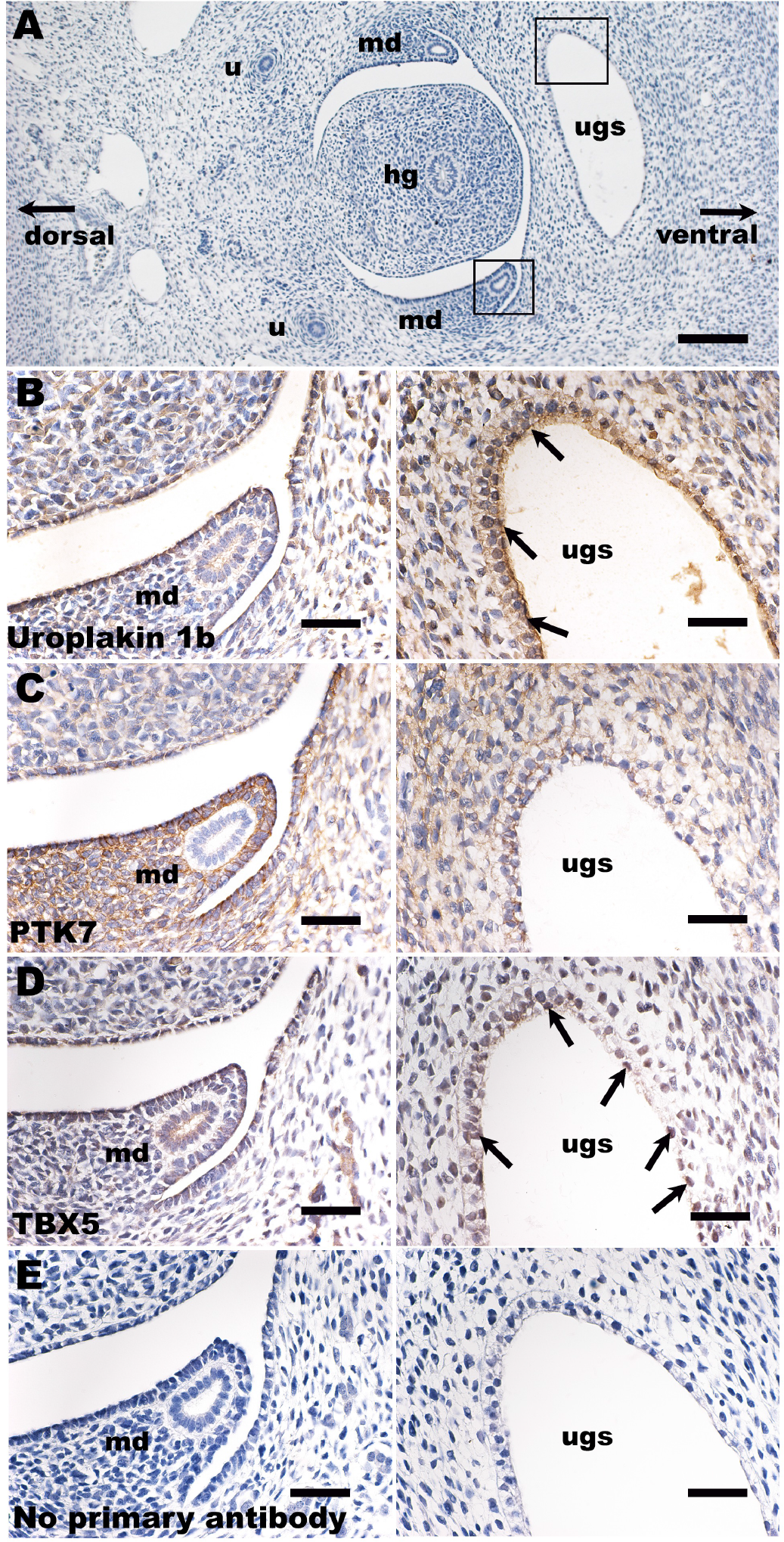
Immunohistochemistry in human embryogenesis. **A**, Overview of transverse section of a normal human embryo seven weeks after fertilization. Key: ugs = urogenital sinus; md = mesonephric duct; hg = hindgut; and u = ureter. The section has been stained with haematoxylin (blue nuclei). Boxes around the ugs and the md mark similar areas depicted under high power in **B-E**. In **B-D**, sections were reacted with primary antibodies, as indicated; in E, the primary antibody was omitted. **B-E** were counterstained with haematoxylin. In **B-E**, the left-hand frame shows the region around the md, while the right-hand frame shows one lateral horn of the ugs. **B.** Uroplakin 1b immunostaining revealed positive signal (brown) in the apical aspect of epithlelia lining the ugs (arrows, right frame), the precursor of the urinary bladder and proximal urethra. Uroplakin 1b was also detected in the flat monolayer of mesothelial cells (left frame) that line the body cavity above the md. **C.** There were strong PTK7 signals (brown cytoplasmic staining) in stromal-like cells around the md (left frame), whereas the epithelia of the duct itself were negative. PTK7 was also detected in a reticular pattern in epithelia lining the ugs (right frame) and in stromal cells near the sinus. **D.** A subset of epithelial cells lining the ugs (right frame) immunostained for TBX5 (brown nuclei; some are arrowed). The mesothelial cells near the md (left frame) were also positive for TBX5. **E.** This negative control section had the primary antibody omitted; no specific (brown) signal was noted. Bar is 400 mm in A, and bars are 100 mm in B-E.

### Monogenic causes of PUV are rare

Having identified two novel gene associations through GWAS, we next aimed to determine whether there was any gene-based enrichment of rare coding variation in PUV cases. Single-variant association tests can be underpowered when variants are rare and collapsing variant data into specific regions or genes can increase power and aid gene discovery. We therefore aggregated rare (gnomAD^24^ allele frequency [AF] < 0.1%), predicted deleterious (protein-truncating, or combined annotation dependent depletion [CADD]^37^ score ≥20) single-nucleotide variants (SNVs) and small indels by gene, comparing the burden between cases and controls on an exome-wide basis. No significant enrichment was detected in any of the 19,364 protein-coding genes analyzed after correction for multiple testing (Fig. 1A). The median number of variants tested per gene was 41 (IQR 47). None of the genes previously associated with congenital bladder outflow obstruction (*BNC2*, *HPSE2, LRIG2, CHRM3, MYOCD*) showed evidence of enrichment. Table S2 lists the genes identified with *P* < 0.01. The absence of gene-based enrichment confirms previous observations that monogenic causes of non-familial PUV are rare.

### Structural variation affecting regulatory elements is enriched

Large, rare CNVs have been identified in patients with PUV using conventional microarrays^19–22^, however high-coverage WGS enables detection of smaller structural variants (SVs) with superior resolution^38, 39^, and allows the identification of balanced rearrangements including inversions. We therefore aimed to detect association with different types of SVs, by comparing the burden of rare (MAF < 0.1%) autosomal SVs on an exome-wide and cis-regulatory element basis.

We first focused our analysis on rare SVs that were potentially gene-disrupting by extracting those that intersected with at least one exon. Although we observed an increased burden of all SV types in cases compared with controls, this only reached statistical significance for inversions (*P*=2.1×10^-3^) when corrected for the multiple SV comparisons performed (Table S3). No difference in SV size between the cohorts was seen. Furthermore, exome-wide gene-based burden analysis did not detect any gene-level enrichment of rare SVs overall or when stratified by type (Table S4), indicating that rare structural variation does not appear to affect any single gene more frequently in PUV than controls.

Given the tightly controlled transcriptional networks that govern embryogenesis we hypothesized that regulatory regions may be preferentially affected by rare structural variation. To investigate this, we identified rare (MAF < 0.1%) autosomal SVs that intersected with 926,535 genome-wide candidate cis-regulatory elements (cCREs) curated by ENCODE^31^. A significant enrichment of cCRE-intersecting SVs was observed for inversions (61.4% vs 47.1%, *P*=1.2×10^-3^) and duplications (78.8% vs 67.5%, *P*=5.0×10^-3^) (Table S3). While the median size of inversions was larger in cases, this was not statistically significant (129kb vs 94kb, *P*=0.12).

To further characterize this enrichment, we repeated the burden analysis stratifying by cCRE subtype (distal enhancer-like signature [dELS], proximal enhancer-like signature [pELS], promoter-like signature [PLS], CTCF-only and DNase-H3K4me3) and demonstrated a consistent signal across all cCREs for inversions (Fig.1B), most significantly affecting CTCF-only elements (49.2% vs 31.7%, *P*=3.1×10^-5^, Table S5). These elements act as chromatin loop anchors suggesting that inversions affecting these regions may potentially alter long-range regulatory mechanisms mediated by chromatin conformation. Duplications affecting pELS elements were also significantly enriched in cases (29.5% vs 16.8%, *P*=2.7×10^-4^).

## Discussion

Using a mixed ancestry whole-genome sequencing approach we have identified the first genetic loci associated with PUV and implicated *TBX5* and *PTK7* in its underlying pathogenesis. Aberrations of mesonephric duct and urogenital sinus maturation have been postulated to be implicated in the pathogenesis of PUV.^6^ Our observations that TBX5 and PTK7 molecules are present during normal human embryogenesis in or around the ducts and the sinus are consistent with the hypothesis that deregulated expression of either may perturb the normal development of these structures. In addition, we detected an enrichment of rare structural variation affecting candidate cis-regulatory elements and demonstrate that monogenic causes of PUV are not a common feature.

The majority of genetic association studies are carried out in individuals of European ancestry, however with next-generation sequencing allowing unbiased variant detection as well as improved statistical methodology to mitigate confounding by population structure, it is widely recognized that increasing ancestral diversity in genetic studies is scientifically and ethically necessary.^27^ GWAS findings have been shown to replicate across populations in a variety of common diseases,^40–47^ suggesting sharing of common causal variants between ancestries despite differences in allele frequencies and effect sizes.^48^ Furthermore, the benefit of combining population groups has been clearly demonstrated in trans-ancestry meta-analyses,^49–51^ where differences in LD structure are specifically utilized to improve the resolution of fine-mapping. Mixed ancestry rare variant analyses are also a useful way to boost power for gene discovery through increased sample size,^52^ with the ‘collapsing’ approach used to aggregate rare variants removing concerns regarding differing allele frequencies across population groups. On this basis we opted to combine individuals regardless of ancestral background and used a generalized mixed model association test with saddlepoint approximation to maximize the signal from the resulting mixed ancestry, case-control imbalanced dataset.

We identified a significant protective effect of rs10774740 (T), highlighting that common variants can contribute to an individual’s risk of a rare disease, as is increasingly being recognized, e.g. for neurodevelopmental disorders.^53^ The effect size and direction were consistent between African, European and South Asian ancestries, despite differences in allele frequency between the population groups. Using experimentally determined chromatin interaction data from mesendoderm cells we mapped this locus to the promoter of the transcription factor *TBX5,* which is known to cause autosomal dominant Holt-Oram syndrome (MIM 142900), characterized by congenital cardiac septal defects and upper-limb anomalies.^54, 55^ No eQTL data were available to determine how rs10774740 might affect *TBX5* expression, however, we show that TBX5 is detected in the urogenital sinus during normal human embryogenesis providing support for its role in lower urinary tract development.

The association of the risk allele (G) with prostate cancer aggressiveness in men and genital prolapse and urinary incontinence in women raises the intriguing possibility that *TBX5,* which shows moderate expression in the adult bladder, is also associated with lower urinary tract phenotypes in adults. Of note, variation in candidate genes associated with other developmental anomalies has also been linked to malignancy in the same organ, e.g., *FOXF1* and VACTERL (vertebral defects, anal atresia, cardiac defects, tracheo-esophageal anomalies, renal anomalies and limb anomalies) with Barrett’s esophagus,^56^ highlighting the common molecular pathways driving both embryogenesis and cancer.

We also identified an association of the rare variant rs144171242 with PUV, located in an intron of *PTK7* and predicted to have regulatory activity in mesendoderm cells. This variant was only seen in European and South Asian groups, suggesting it arose after migration from Africa. The inclusion of South Asian individuals, in whom the effect size of rs144171242 is larger, increased our power to detect association which was not genome-wide significant in the European-only analysis. PTK7 (protein tyrosine kinase 7) is an evolutionarily conserved transmembrane receptor required for vertebrate embryonic patterning and morphogenesis, and a key regulator of planar cell polarity (PCP) via the non-canonical Wnt pathway.^57^ The PCP pathway is critical for determining the orientation of cells in the plane of an epithelium, regulating a process called convergent extension whereby cells intercalate by converging in one axis and elongating in the perpendicular axis. Altered expression of *PTK7* was initially observed in cancer,^58, 59^ but rare missense variants in *PTK7* have since been linked to neural tube defects^60, 61^ and scoliosis^62^ in both humans and animal models, confirming a role in embryonic development. In our study, PTK7 was detected in stromal-like cells surrounding the mesonephric ducts and the urogenital sinus indicating it is present during normal embryonic development. Interestingly, mesoderm-specific conditional deletion of *Ptk7* in mice has been shown to affect convergent extension and tubular morphogenesis of the mesonephric duct at E18.5, leading to male sterility.^63^ Whether similar disruption in mesonephric duct morphogenesis is seen at E14 (corresponding to development of the urethra) remains to be seen but may provide further insights into the biological mechanisms underpinning PUV.

Rare CNVs have been associated with neurodevelopmental disorders^64–68^ and congenital malformations^69, 70^ and recently shown to be enriched in patients with kidney and urinary tract anomalies^22^. However, our study, consistent with a previous microarray-based study by Verbitsky *et al.,*^22^ did not identify an increased burden of CNVs in individuals with PUV. We observed a higher number of rare, exonic CNVs than Verbitsky *et al.*^22^ (82.6% vs 32.6%), most likely reflecting the increased sensitivity and resolution of WGS for SV detection as well as the difference in size threshold for inclusion (> 10kb compared to >100kb). Importantly, none of the CNVs recurrently affected a particular gene which, in combination with the lack of gene-based enrichment seen in our rare SNV/indel analysis, confirms previous observations that monogenic causes of PUV are rare, although our sample size would be underpowered to detect significant genetic heterogeneity.

Intriguingly, we demonstrated an enrichment of rare inversions affecting cCREs. Current understanding of the functional relevance of inversions is limited as the balanced nature and location of breakpoints within complex repeat regions make detection challenging.^71^ Although usually considered neutral, inversions can directly disrupt coding sequences or regulatory elements, as well as predispose to other SVs, and have been associated with hemophilia A,^72^ Hunter syndrome,^73^ neurodegenerative^74^ and autoimmune disease.^75, 76^ The strongest signal we observed was for inversions affecting CTCF-only regions, potentially implicating disrupted chromatin looping in the underlying pathogenesis of PUV. The enrichment of rare inversions affecting cCREs raises the interesting possibility that non-specific perturbation of long-range regulatory networks or TADs could manifest as PUV, perhaps due to sensitivity of integration of the mesonephric duct into the posterior urethra to even minor abnormalities of gene expression.

This study has several strengths. WGS enables ancestry independent variant detection, uniform genome-wide coverage, improved SV resolution and detection, as well as direct sequencing of underlying causal variants. Using case-control data from >20,000 individuals sequenced on the same platform also minimizes confounding by technical artefacts. Inclusion of diverse ancestries increased our power to detect both novel associations and the underlying causal variant, with the lack of genomic inflation and subsequent replication indicating these associations are robust. Furthermore, we integrated GWAS, epigenomic and chromatin interaction data to ascertain the functional relevance of loci and identify biologically plausible genes.

A limitation of this study is its relatively low statistical power to detect associations with small effects and future meta-analyses with larger cohorts are necessary to identify additional loci. Furthermore, while WGS offers improved SV resolution over microarrays, false positives may occur and are dependent on the SV calling algorithm used. Ideally, long-read sequencing and independent validation would be used to provide more comprehensive SV detection, especially of larger variants in complex, repetitive and GC-rich regions. Finally, although we have assessed the relevance of the associated loci using bioinformatic approaches and shown that publicly available and our own experimental data support the association, the biological mechanisms linking these genes with PUV have yet to be elucidated.

To our knowledge, this is the first study to utilize mixed ancestry WGS for association testing in a rare disease. Combining WGS data across ancestries increased power, revealed two novel loci for PUV and identified the likely causal variants through enhanced fine-mapping. Finally, integration of functional genomic and experimental data implicated *TBX5* and *PTK7* in the pathogenesis of PUV, an important but poorly understood disorder.

## Methods

### The 100,000 Genomes Project (100KGP)

The Genomics England dataset^23^ (v10) consists of whole-genome sequencing (WGS) data, clinical phenotypes encoded using Human Phenotype Ontology^77^ (HPO) codes, and retrospective and prospectively ascertained National Health Service (NHS) hospital records for 89,139 individuals recruited with cancer, rare disease, and their unaffected relatives. Ethical approval for the 100KGP was granted by the Research Ethics Committee for East of England – Cambridge South (REC Ref 14/EE/1112). Fig. S1 details the study workflow.

Cases were recruited from 13 NHS Genomic Medicine Centers across the UK as part of the 100KGP ‘Congenital anomalies of the kidneys and urinary tract (CAKUT)’ cohort with the following inclusion criteria: CAKUT with syndromic manifestations in other organ systems; isolated CAKUT with a first-degree relative with CAKUT or unexplained CKD; multiple distinct renal/urinary tract anomalies; CAKUT with unexplained end-stage kidney disease before the age of 25 years. Those with a clinical or molecular diagnosis of ADPKD or ARPKD, or who had a known genetic or chromosomal abnormality were excluded. 136 male individuals with a diagnosis of posterior urethral valves (PUV) were identified using the HPO term “HP:0010957 congenital posterior urethral valve”.

All cases underwent assessment via the clinical interpretation arm of the 100KGP to determine a molecular diagnosis. This process involved the examination of protein-truncating and missense variants from an expert-curated panel of 57 CAKUT-associated genes followed by multi-disciplinary review and application of ACMG^78^ criteria to determine pathogenicity. CNVs affecting the 17q12 region (ISCA-37432Loss), which includes *HNF1B*, were also assessed. No pathogenic/likely pathogenic variants were identified in genes previously associated with congenital bladder outflow obstruction (*HPSE2, LRIG2, CHRM3, MYOCD, BNC2*). Two pathogenic/likely pathogenic variants affecting the 17q12 locus and *FOXC1* were identified in two individuals, but these were not deemed to be causal for PUV (see Supplemental Note).

The control cohort consisted of 27,660 unaffected relatives of non-renal rare disease participants, excluding those with HPO terms and/or hospital episode statistics (HES) data consistent with kidney disease or failure. By utilizing a case-control cohort sequenced on the same platform, we aimed to minimize confounding by technical artefacts.

### DNA preparation and extraction

99% of DNA samples were extracted from blood and prepared using EDTA, with the remaining 1% sourced from saliva, tissue, and fibroblasts. Samples underwent quality control assessment based on concentration, volume, purity, and degradation. Libraries were prepared using the Illumina TruSeq DNA PCR-Free High Throughput Sample Preparation kit or the Illumina TruSeq Nano High Throughput Sample Preparation kit.

### Whole-genome sequencing, alignment, and variant calling

Samples were sequenced with 150bp paired-end reads using an Illumina HiSeq X and processed on the Illumina North Star Version 4 Whole Genome Sequencing Workflow (NSV4, version 2.6.53.23), comprising the iSAAC Aligner (version 03.16.02.19) and Starling Small Variant Caller (version 2.4.7). Samples were aligned to the Homo Sapiens NCBI GRCh38 assembly. Alignments had to cover ≥ 95% of 15X with mapping quality > 10 for samples to be retained. Samples achieved a mean of 97.4% coverage at 15X with a median genome-wide coverage of 39X. Samples with <2% cross-contamination as determined by the VerifyBamID algorithm were kept. Copy number and structural variant (>50bp) calling was performed using CANVAS^79^ (version 1.3.1) and MANTA^80^ (version 0.28.0) respectively. CANVAS determines coverage and minor allele frequencies (MAF) to assign copy number (>10kb) whereas MANTA combines paired and split-read algorithms to detect structural variants (< 10kb).

### gVCF annotation and variant-level quality control

gVCFs were aggregated using gvcfgenotyper (Illumina, version: 2019.02.26) with variants normalized and multi-allelic variants decomposed using vt^81^ (version 0.57721). Variants were retained if they passed the following filters: missingness ≤ 5%, median depth ≥ 10, median GQ ≥ 15, percentage of heterozygous calls not showing significant allele imbalance for reads supporting the reference and alternate alleles (ABratio) ≥ 25%, percentage of complete sites (completeGTRatio) ≥ 50% and *P* value for deviations from Hardy-Weinberg equilibrium (HWE) in unrelated samples of inferred European ancestry ≥ 1×10^-5^. Male and female subsets were analyzed separately for sex chromosome quality control. Per-variant minor allele count (MAC) was calculated across the case-control cohort. Annotation was performed using Variant Effect Predictor^82^ (VEP, version 98.2) including CADD^37^ (version 1.5), and allele frequencies from publicly available databases including gnomAD^24^ (version 3) and TOPMed^83^ (Freeze 5). Variants were filtered using bcftools^84^ (version 1.11).

### Relatedness estimation and principal components analysis

A set of 127,747 high quality autosomal LD-pruned biallelic single nucleotide variants (SNVs) with MAF > 1% was generated using PLINK^85^ (v1.9). SNVs were included if they met all the following criteria: missingness < 1%, median GQ ≥ 30, median depth ≥ 30, AB Ratio ≥ 0.9, completeness ≥ 0.9. Ambiguous SNVs (AC or GT) and those in a region of long-range high LD were excluded. LD pruning was carried out using an r^2^ threshold of 0.1 and window of 500kb. SNVs out of HWE in any of the AFR, EAS, EUR or SAS 1000 Genomes populations were removed (pHWE < 1 ×10^-5^). Using this variant set, a pairwise kinship matrix was generated using the PLINK2^86^ implementation of the KING-Robust algorithm^87^ and a subset of unrelated samples was ascertained using a kinship coefficient threshold of 0.0884 (2nd degree relationships). Two cases and 1,354 controls were found to be related by this method and were removed, leaving 134 cases and 26,306 controls. Ten principal components were generated using PLINK2^86^ for ancestry-matching and for use as covariates in the association analyses.

### Ancestry-matching of cases and controls

Given the mixed-ancestry composition of the cohort we employed a case-control ancestry-matching algorithm to optimize genomic similarity and minimize the effects of population structure. A custom R script (see Code availability) was used to match cases to controls within a distance threshold calculated using the top ten principal components weighted by the percentage of genetic variation explained by each component (Fig. S2). Only controls within a user-defined specified distance of a case were included with each case having to match a minimum of two controls to be included in the final cohort. A total of two cases and 2,579 controls were excluded using this approach, leaving 132 cases and 23,727 controls for further analysis.

### GWAS

Genome-wide single variant association analysis was carried out using the R package SAIGE^88^ (version 0.42.1) which uses a generalized logistic mixed model (GLMM) to account for population stratification, and is recommended for use in mixed-ancestry cohorts.^27^ 2,000 randomly selected high-quality, autosomal, bi-allelic, LD-pruned SNVs with MAF > 5% were used to generate a genetic relationship matrix and fit the null GLMM. Sex and the top ten principal components were used as fixed effects. SNVs and indels with MAF > 0.1% and that passed the following quality control filters were retained: MAC ≥ 20, missingness < 1%, HWE *P* > 10 and differential missingness *P* > 10^-5^. A score test^89^ for association was performed for 17,091,503 variants (mixed-ancestry GWAS) and 15,447,192 variants (European-only GWAS). When case-control ratios are unbalanced, as in our study (1:180), type 1 error rates are inflated because the asymptotic assumptions of logistic regression are invalidated. Like SAIGE-GENE, SAIGE employs a saddlepoint approximation^90^ to calibrate score test statistics and obtain more accurate *P* values than the normal distribution.

At each of the genome-wide significant loci we used SAIGE to perform a) conditional analysis to identify secondary independent associations and b) high resolution single variant analysis using all variants with MAC ≥ 3 to ascertain whether the observed signal was being driven by rare variation. Epistasis between the lead variants was assessed using logistic regression in PLINK^85^ (version 1.9). One limitation of SAIGE is that the betas estimated from score tests can be biased at low MACs and therefore odds ratios for variants with MAF < 1% were calculated separately using allele counts in R. The R packages qqman^91^ and GWASTools^92^ were used to create Manhattan and Q-Q plots, and LocusZoom^93^ to visualize regions of interest.

### Replication

The replication cohort consisted of 398 individuals with PUV; 336 recruited from Poland and Germany as part of the CaRE for LUTO (Cause and Risk Evaluation for Lower Urinary Tract Obstruction) Study, and 62 from Manchester, UK. None of the individuals had been recruited to the 100KGP. All were of self-reported European ancestry. KASP (Kompetitive Allele-Specific PCR) genotyping of the lead variants at the top four loci using a threshold of *P* < 5×10^-7^ was carried out: rs10774740 at 12q24.21, rs144171242 at 6p21.1, rs1471950716 at 10q11.21, rs199975325 at 14q21.1. The peri-centromeric location of rs1471950716 at 10q11.21 caused the genotyping assay to fail and another variant with evidence of association (rs137855548; *P*=1.46×10^-6^) was used instead. The control cohort consisted of 10,804 genetically determined European individuals recruited to the cancer arm of the 100KGP, excluding those with kidney, bladder, prostate, or childhood malignancy. Allele counts at each variant were compared between cases and controls using a one-sided Cochran-Armitage trend test. A Bonferroni-corrected *P* < 0.0125 (0.05/4) was used to adjust for the number of loci tested. Power to detect or refute association at each locus was calculated as > 0.9.

### Power

Statistical power for single-variant association under an additive model for the discovery and replication cohorts was calculated using the R package (genpwr).^94^ Fig. S8 shows the power calculations for the mixed-ancestry GWAS at varying allele frequencies and odds ratios.

### Bayesian fine-mapping

We applied PAINTOR^25^ (v3.1), a statistical fine-mapping method which uses an empirical Bayes prior to integrate functional annotation data, linkage disequilibrium (LD) patterns and strength of association to estimate the posterior probability (PP) of a variant being causal. Variants at each genome-wide significant locus with *P* < 0.05 were extracted. Z-scores were calculated as effect size (β) divided by standard error.

LD matrices of pairwise correlation coefficients were derived using EUR 1,000 Genomes^95^ (Phase 3) imputed data as a reference, excluding variants with ambiguous alleles (A/T or G/C). Each locus was intersected with the following functional annotations downloaded using UCSC Table Browser^28^: GENCODE^96^ (v29) transcripts (wgEncodeGencodeBasicV29, updated 2019-02-15), PhastCons^97^ (phastConsElements100way, updated 2015-05-08), ENCODE^31^ cCREs (encodeCcreCombined, updated 2020-05-20), transcription factor binding clusters (encRegTfbsClustered, updated 2019-05-16), DNase I hypersensitivity clusters (wgEncodeRegDnaseClustered, updated 2019-01-08) and H1 Human embryonic stem cell Hi-C data (h1hescInsitu from Krietenstein *et al.*, 2020^98^). A total of 351 variants at 12q24.21 and 166 variants at 6p21.1 were analyzed under the assumption of one causal variant per locus.

### Functional annotation

To explore the functional relevance of the prioritized variants we used FUMA^29^ (v1.3.6a) to annotate the genome-wide significant loci. This web-based tool integrates functional gene consequences from ANNOVAR^99^, CADD^37^ scores to predict deleteriousness, RegulomeDB score to indicate potential regulatory function^100^ and 15-core chromatin state (predicted by ChromHMM for 127 tissue/cell types)^101^ representing accessibility of genomic regions. Positional mapping (where a variant is physically located within a 10kb window of a gene), GTEx (v8) eQTL data^102^ (using cis-eQTLs to map variants to genes up to 1Mb apart) and Hi-C data to detect long-range 3D chromatin interactions is used to prioritize genes that are likely to be affected by variants of interest. GWAS summary statistics were used as input with genomic positions converted to GRCh37 using the UCSC^28^ liftOver tool.

In addition, we intersected prioritized variants with the following epigenomic datasets from male H1-BMP4 derived mesendoderm cultured cells generated by the ENCODE Project^31^ and Roadmap Epigenomics^34^ Consortia using the UCSC Genome Browser^28^: ENCFF918FRW_ENCFF748XLQ_ENCFF313DOD (cCREs, GRCh38); ENCFF918FRW_ENCFF748XLQ_ENCFF313DOD_ENCFF313DOD (H3K27ac ChIP-seq, GRCh38); ENCFF918FRW_ENCFF748XLQ_ENCFF313DOD_ENCFF748XLQ (H3K4me3 ChIP-seq, GRCh38); ENCFF918FRW_ENCFF748XLQ_ENCFF313DOD_ENCFF918FRW (DNase-seq, GRCh38); E004 H1 BMP4 Derived Mesendoderm Cultured Cells ImputedHMM (hg19). Hi-C interactions from H1 mesendoderm cells^32^ and topologically associated domains (TADs) were visualized with the 3D Interaction Viewer and Database (see Web resources).

### Gene and gene-set analysis

MAGMA^103^ (v1.6) was used to test the joint association of all variants within a particular gene or gene-set using the GWAS summary statistics. Aggregation of variants increases power to detect multiple weaker associations and can test for association with specific biological or functional pathways. MAGMA uses a multiple regression approach to account for LD between variants, using a reference panel derived from 10,000 Europeans in the UK Biobank (release 2b). Variants from the GWAS were assigned to 18,757 protein coding genes (Ensembl build 85) with genome-wide significance defined as *P*=2.67×10^-6^ (0.05/18,757). Competitive gene-set analysis was then performed for 5,497 curated gene sets and 9,986 Gene Ontology (GO) terms from MsigDB^104^ (version 7.0) using the results of the gene analysis. Competitive analysis tests whether the joint association of genes in a gene-set is stronger than a randomly selected set of similarly sized genes. Bonferroni correction was applied for the total number of tested gene sets (*P*=0.05/15,483=3.23×10^-6^).

### Identification of TFBS

The JASPAR 2020^30^ CORE collection track (UCSC Genome Browser^28^, updated 2019-10-13) was utilized to identify significant (*P* < 10^-4^) predicted TFBS that might intersect with the lead variants. The JASPAR database consists of manually curated, non-redundant, experimentally defined transcription factor binding profiles for 746 vertebrates, of which 637 are associated with human transcription factors with known DNA-binding profiles. Sequence logos based on position weight matrices of the DNA binding motifs were downloaded from JASPAR 2020.^30^

### GWAS and PheWAS associations

The NHGRI-EBI GWAS Catalog^35^ and PheWAS data from the UK Biobank (see Web resources) were interrogated to determine known associations of the lead variants. Summary statistics were downloaded from the NHGRI-EBI GWAS Catalog^35^ for study GCST002890^36^ on 17/03/2021. PheWAS statistics were generated using imputed data from White British participants in the UK Biobank using SAIGE, adjusting for genetic relatedness, sex, birth year and the first four principal components.

### Immunohistochemistry

Human embryonic tissues, collected after maternal consent and ethical approval (REC18/NE/0290), were sourced from the Medical Research Council and Wellcome Trust Human Developmental Biology Resource (https://www.hdbr.org/). Tissues sections were immunostained, as we described previously.^11^ Sections were immunostained with the following primary antibodies: TBX5 (https://www.abcam.com/tbx5-antibody-ab223760.html) raised in rabbit; PTK7 (https://www.thermofisher.com/antibody/product/PTK7-Antibody-Polyclonal/PA5-82070) raised in rabbit; and uroplakin 1B (https://www.abcam.com/uroplakin-ibupib-antibody-upk1b3081-ab263454.html) raised in mouse. Primary antibodies were detected with appropriate second antibodies and signals generated with a peroxidase-based system.

### Aggregate rare coding variant analysis

Single variant association testing is underpowered when variants are rare and a collapsing approach which aggregates variants by gene can be adopted to boost power. We extracted coding SNVs and indels with MAF < 0.1% in gnomAD,^24^ annotated with one of the following: missense, in-frame insertion, in-frame deletion, start loss, stop gain, frameshift, splice donor or splice acceptor. Variants were further filtered by CADD^37^ (v1.5) score using a threshold of ≥20 corresponding to the top 1% of all predicted deleterious variants in the genome. Variants meeting the following quality control filters were retained: MAC ≤ 20, median site-wide depth in non-missing samples > 20 and median GQ ≥ 30. Sample-level QC metrics for each site were set to minimum depth per sample of 10, minimum GQ per sample of 20 and

ABratio *P* value > 0.001. Variants with significantly different missingness between cases and controls (*P*<10^-5^) or >5% missingness overall were excluded. We employed SAIGE-GENE^105^ (v0.42.1) to ascertain whether rare coding variation was enriched in cases on a per-gene basis exome-wide. SAIGE-GENE utilizes a generalized mixed-model to correct for population stratification and cryptic relatedness as well as a saddlepoint approximation and efficient resampling adjustment to account for the inflated type 1 error rates seen with unbalanced case-control ratios. It combines single-variant score statistics and their covariance estimate to perform SKAT-O^106^ gene-based association testing, upweighting rarer variants using the beta(1,25) weights option. SKAT-O^106^ is a combination of a traditional burden and variance-component test and provides robust power when the underlying genetic architecture is unknown. Sex and the top ten principal components were included as fixed effects when fitting the null model. A Bonferroni adjusted *P* value of 2.58×10^-6^ (0.05/19,364 genes) was used to determine the exome-wide significance threshold.

### Structural variant analysis

Structural variants (>50bp) that intersect by a minimum of 1bp with a) at least one exon (GENCODE^96^; version 29) or b) an ENCODE^31^ candidate cis-regulatory element (cCRE) were extracted using BEDTools^107^ (version 2.27.1). Variants were retained if they fulfilled the following quality filters: Q-score ≥ Q10 (CANVAS) or QUAL ≥ 20, GQ ≥ 15, and MaxMQ0Frac < 0.4 (MANTA). Variants without paired read support, inconsistent ploidy, or depth >3x the mean chromosome depth near breakends were excluded.

ENCODE^31^ cCREs are 150-350bp consensus sites of chromatin accessibility (DNase hypersensitivity sites) with high H3K4me3, high H3K27ac, and/or high CTCF signal in at least one biosample. A list of 926,535 cCREs encoded by 7.9% of the human genome was downloaded from UCSC Table Browser using the encodeCcreCombined track (updated 20/05/2020). This includes ∼668,000 distal enhancer-like signature (dELS) elements, ∼142,000 proximal enhancer-like signature (pELS) elements, ∼57,000 CTCF-only elements, ∼35,000 promoter-like signature (PLS) and ∼26,000 DNase-H3K4me3 elements (promoter-like signals > 200bp from a transcription start site).

Variants were separated and filtered by SV type (deletion, duplication, CNV, inversion); those with a minimum 70% reciprocal overlap with common SVs from a) dbVar^108^ or b) 12,234 cancer patients from the 100KGP were removed. The dbVar NCBI curated dataset of SVs (nstd186) contains variant calls from studies with at least 100 samples and AF > 1% in at least one population, including gnomAD^24^, 1000 Genomes (Phase 3)^95^ and DECIPHER.^67^ To create a dataset of common SVs from the 100KGP cancer cohort, variants were merged using SURVIVOR^109^ (v1.0.7), allowing a maximum distance of 300bp between pairwise breakpoints, and those with AF > 0.1% retained. After removal of overlapping common variants, SVs in the case-control cohort were filtered to keep those with AF < 0.1% and aggregated across 19,907 autosomal protein-coding genes and five cCRE types. Exome-wide gene-based and genome-wide cCRE-based burden analysis was carried out using custom R scripts. The burden of rare autosomal SVs in cases and controls was enumerated by comparing the number of individuals with ≥ Fisher’s Exact test. The Wilcoxon-Mann-Whitney test was used to compare median SV size. Bonferroni adjustment for the number of genes (*P*=0.05/19,907=2.5×10^-6^) and cCRE/SV combinations (*P*=0.05/20=2.5×10^-3^) tested was applied.

## Data and code availability

Genomic and phenotype data from participants recruited to the 100KGP can be accessed by application to Genomics England Ltd (https://www.genomicsengland.co.uk/about-gecip/joining-research-community/).

Code for the case-control ancestry-matching algorithm can be found at https://github.com/APLevine/PCA_Matching.

## Data Availability

Genomic and phenotype data from participants recruited to the 100,000 Genomes Project can be accessed by application to Genomics England Ltd (https://www.genomicsengland.co.uk/about-gecip/joining-research-community/).

## Acknowledgements

This research was made possible through access to the data and findings generated by the 100,000 Genomes Project. The 100,000 Genomes Project is managed by Genomics England Limited (a wholly owned company of the Department of Health and Social Care). The 100,000 Genomes Project is funded by the National Institute for Health Research and NHS England. The Wellcome Trust, Cancer Research UK and the Medical Research Council have also funded research infrastructure. The 100,000 Genomes Project uses data provided by patients and collected by the National Health Service as part of their care and support. The authors gratefully acknowledge the participation of the patients and their families recruited to the 100,000 Genomes Project. The authors also gratefully acknowledge the participation of the patients and their families in the replication study, the majority of whom were recruited via the CaRE for LUTO Study.

MMYC is funded by a Kidney Research UK Clinical Research Training Fellowship (TF_004_20161125). OSA is funded by an MRC Clinical Research Training Fellowship. ACH is supported by a BONFOR-Gerok Grant. WGN, ASW, and GMB are supported by Kidney Research UK (Paed_RP_002_20190925). ASW receives funding from the Medical Research Council (MR/T016809/1). APL is supported by an NIHR Academic Clinical Lectureship. DPG is supported by the St Peter’s Trust for Kidney, Bladder and Prostate Research.

## Declaration of interests

The authors declare no competing interests.

## Supplemental Data

**Supplemental Appendix**: Genomics England Research Consortium

**Supplemental Note:** Pathogenic/likely pathogenic variants identified in PUV cases

**Figure S1 | Study workflow.** The flowchart shows the number of samples included at each stage of filtering, the analytical strategies employed and the main findings (blue boxes). PUV, posterior urethral valves; MAF, minor allele frequency; GWAS, genome-wide association study; EUR, European; cCRE, candidate cis-regulatory element.

**Figure S2 | Ancestry matching.** Principal component analysis showing the first eight principal components for matched cases (blue) and controls (black) and unmatched cases (orange) and controls (grey). Two cases and 2,579 controls were excluded from downstream analyses.

**Figure S3 | Q-Q plot for mixed-ancestry GWAS.** Quantile-quantile (Q-Q) plot displaying the observed versus the expected –log_10_(*P*) for each variant tested. The grey shaded area represents the 95% confidence interval of the null distribution.

**Figure S4 | European GWAS.** A genome-wide single-variant association study was carried out in 88 cases and 17,993 controls for 15,447,192 variants with MAF > 0.1%. All cases and controls had genetically determined European ancestry. **A**, Manhattan plot with chromosomal position (GRCh38) denoted along the *x* axis and strength of association using a –log_10_(*P*) scale on the *y* axis. Each dot represents a variant. The red line indicates the Bonferroni adjusted threshold for genome-wide significance (*P* < 5×10^-8^). The two genome-wide significant loci from the mixed ancestry GWAS are labelled. **B**, Quantile-Quantile (Q-Q) plot displaying the observed versus the expected –log_10_(*P*) for each variant tested. The grey shaded area represents the 95% confidence interval of the null distribution.

**Figure S5 | Correlation between mixed-ancestry and European GWAS.** Comparison of **A**, −log_10_(*P)* and **B**, BETA from the mixed-ancestry and European-only ancestry GWAS. All variants with *P* < 10^-5^ in both cohorts are shown. The shaded grey area represents the 95% confidence interval.

**Figure S6 | Comparison of ancestry-specific odds ratios and gnomAD allele frequencies.** GWAS per-ancestry odds ratios for **A**, rs10774740 and **B**, rs144171242. Comparison of population minor allele frequencies from gnomAD with case and control allele frequencies from our data for **C**, rs10774740 and **D**, rs144171242. Error bars represent 95% confidence intervals. The dashed lines indicate the minor allele frequency observed in cases (orange) and controls (blue) with the shaded areas indicating 95% confidence interval for each group. No data was available for rs10774740 in the South Asian population. AFR, African/African-American; AKJ, Ashkenazi Jewish; EAS, East Asian; EUR, European (non-Finnish); FIN, European (Finnish); LAT, Latino/Admixed-American; OTH, Other; SAS, South Asian.

**Figure S7 | Linkage disequilibrium (LD) for reference populations in the 1000 Genomes Project.** LD plots for 503 European (EUR), 489 South Asian (SAS) and 661 African (AFR) ancestry individuals from the 1000 Genomes Project (Phase 3). Haploview (v4.2) was used to compute pairwise LD statistics (r^2^) between variants for each population. The darker the shading, the higher the LD between variants. Black outlined triangles indicate haploblocks. **A**, LD plot for chr12:114,641,202-114,691,202 (GRCh37) with the position of the lead variant rs10774740 represented by a green arrow; **B**, LD plot for chr6:43,063,094-43,113,094 (GRCh37) with the position of the lead variant rs144171242 represented by a green arrow. rs144171242 was not seen in the AFR population group.

**Figure S8 | GWAS power calculation.** Power calculations were performed at various minor allele frequencies (MAF) using 132 cases and 23,727 controls under an additive genetic model to achieve genome-wide significance of *P*<5×10^-8^.

**Table S1. Clinical characteristics and genetic ancestry**. PUV, posterior urethral valves; PCA, principal components analysis; EUR, European; SAS, South Asian; AFR, African; AMR, Latino/Admixed American; VUR, vesico-ureteral reflux; UTI, urinary tract infection; ESRD, end-stage renal disease.

**Table S2. Exome-wide rare SNV/indel analysis**. Results from SAIGE-GENE aggregate rare (MAF < 0.1%) coding variant association. Gene name and Ensembl identifier listed for all genes with *P* < 0.01. See supplemental data.

**Table S3. Structural variant analysis.** The burden of rare autosomal structural variants intersecting with a) at least one exon or b) a cis-regulatory element was compared between 132 cases and 23,727 controls. cCRE, candidate cis-regulatory element; PUV, posterior urethral valves; CNV, copy number variant; DEL, deletion; DUP, duplication; INV, inversion; OR, odds ratio; CI, 95% confidence interval, IQR, interquartile range.

**Table S5. Structural variant cCRE analysis.** The burden of rare autosomal structural variants intersecting with each cis-regulatory element type was compared between 132 cases and 23,727 controls. cCRE, candidate cis-regulatory element; CNV, copy number variant; DEL, deletion; DUP, duplication; INV, inversion; dELS, distal enhancer-like signature; pELS, proximal enhancer-like signature; PLS, promoter-like signature; OR, odds ratio; CI, 95% confidence interval.

**Table S6. Mixed ancestry GWAS association statistics.** Summary statistics for all variants with *P* < 10^-5^. Allele 2 refers to the effect allele. Allele 1 is the other allele. CHR, chromosome; POS, genomic position with reference to GRCh38; SE, standard error; AF, allele frequency. See supplemental data.

**Table S7. GWAS gene-based analysis.** MAGMA was used to assess the joint effect of common and low-frequency variants across genes. Genes with *P* < 0.01 are listed. CHR, chromosome; START and STOP denote the genomic position with reference to GRCh37; NSNPS; the number of variants aggregated for each gene. See supplemental data.

**Table S8. GWAS pathway analysis.** MAGMA was used to assess the joint effect of common and low-frequency variants across different biological pathways. Pathways with *P* < 0.05 are listed. NGENES; the number of genes aggregated across each pathway; SE, standard error; GO, gene ontology. See supplemental data.

**Table S9. Replication study.** The lead variants at the top four loci with *P* < 5×10^-7^ were genotyped in an independent European cohort of 398 PUV cases and 10,804 controls. *P* values in the replication cohort were calculated using a one-sided Cochran Armitage Trend test. OR, odds ratio; CI, 95% confidence interval.

**Table S10. Comparison of mixed ancestry and European GWAS association statistics.** The lead variants at the top four loci with *P* < 5×10^-7^ are shown. OR, odds ratio; CI, 95% confidence interval.

## Notes

### Competing Interest Statement

The authors have declared no competing interest.

### Author Declarations

Ethical approval for The 100,000 Genomes Project was granted by the Research Ethics Committee for East of England Cambridge South (REC Ref 14/EE/1112). Human embryonic tissues, collected after maternal consent, were sourced from the Medical Research Council and Wellcome Trust Human Developmental Biology Resource (http://www.hdbr.org/). Ethical approval for the HDBR was gained from the Research Ethics Committee for North East - Newcastle (REC18/NE/0290).

### Summary of Updates

This version was revised to add immunohistochemistry data to demonstrate the expression of TBX5 and PTK7 in a human embryo; Figure 5 added; author added and order updated.

