## Supplemental Material for "Mixed ancestry analysis of whole-genome sequencing identifies *TBX5* and *PTK7* as susceptibility genes for posterior urethral valves"

### Supplemental Appendix

#### Genomics England Research Consortium

John C. Ambrose[1]; Prabhu Arumugam[1]; Roel Bevers[1]; Marta Bleda[1]; Freya Boardman-Pretty[1,2]; Christopher R. Boustred[1]; Helen Brittain[1]; Mark J. Caulfield[1,2]; Georgia C. Chan[1]; Greg Elgar[1,2]; Tom Fowler[1]; Adam Giess[1]; Angela Hamblin[1]; Shirley Henderson[1,2]; Tim J. P. Hubbard[1]; Rob Jackson[1]; Louise J. Jones[1,2]; Dalia Kasperaviciute[1,2]; Melis Kayikci[1]; Athanasios Kousathanas[1]; Lea Lahnstein[1]; Sarah E. A. Leigh[1]; Ivonne U. S. Leong[1]; Javier F. Lopez[1]; Fiona Maleady-Crowe[1]; Meriel McEntagart[1]; Federico Minneci[1]; LoukasMoutsianas[1,2]; Michael Mueller[1,2]; Nirupa Murugaesu[1]; Anna C.Need[1,2]; Peter O'Donovan[1]; Chris A.Odhams[1]; Christine Patch[1,2]; Mariana Buongermino Pereira[1]; Daniel Perez-Gil[1]; John Pullinger[1]; Tahrima Rahim[1]; Augusto Rendon[1]; Tim Rogers[1]; Kevin Savage[1]; Kushmita Sawant[1]; Richard H. Scott[1]; Afshan Siddiq[1]; Alexander Sieghart[1]; Samuel C. Smith[1]; Alona Sosinsky[1,2]; Alexander Stuckey[1]; Mélanie Tanguy[1]; Ana Lisa Taylor Tavares[1]; Ellen R. A. Thomas[1,2]; Simon R. Thompson[1]; Arianna Tucci[1,2]; Matthew J. Welland[1]; Eleanor Williams[1]; Katarzyna Witkowska[1,2]; Suzanne M.Wood[1,2].

1. Genomics England, London, UK
2. William Harvey Research Institute, Queen Mary University of London, London, EC1M 6BQ, UK.

#### Supplemental Note

**Pathogenic/likely pathogenic variants identified in PUV cases**

One individual was heterozygous for a pathogenic 1.5Mb 17q12 deletion (affecting *HNF1B*) which has previously been associated with autosomal dominant renal cysts and diabetes syndrome (RCAD, OMIM 137920). Review of available clinical information revealed the presence of renal cysts, hypomagnesemia and hypokalemia, which is consistent with *HNF1B*-related disease. There was no reported family history which is often seen since ~50% of whole gene deletions are *de novo* and there is variable expressivity. Identification of this variant was reported back to the clinical team for validation and to inform surveillance for additional manifestations (e.g., diabetes, hyperuricemia, autism) and screening of family members. Review of hospital records confirmed that this individual had undergone endoscopic valve ablation for PUV as an infant and therefore had two separate diagnoses with the 17q12 deletion not determined as causal for PUV.

Another individual with a diagnosis of PUV, ocular hypertension, neurodevelopmental delay and autistic behavior was found to be heterozygous for a likely pathogenic missense variant *FOXC1:*c.379C>T; p.(Arg127Cys). This variant has been previously reported in individuals with pediatric glaucoma (PMID 28979898) and heterozygous *FOXC1* variants have been identified in individuals with syndromic CAKUT, one of whom had PUV (PMID 32475988). After multi-disciplinary review of the available evidence, this variant was not thought to fully explain the patient's phenotype.

#### Figure S1

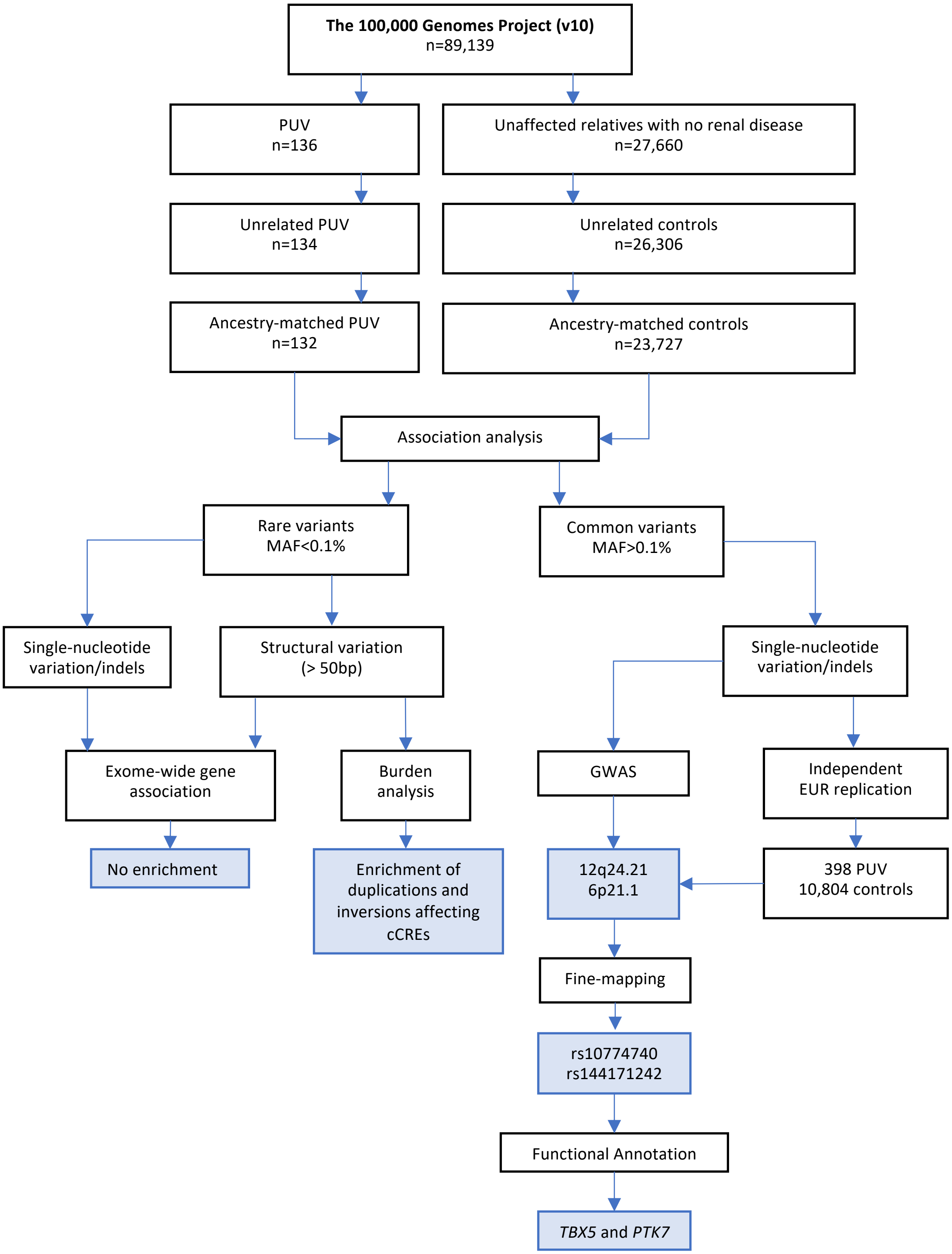


**Figure S1 | Study workflow.** The flowchart shows the number of samples included at each stage of filtering, the analytical strategies employed and the main findings (blue boxes). PUV, posterior urethral valves; MAF, minor allele frequency; GWAS, genome-wide association study; EUR, European; cCRE, candidate cis-regulatory element.

### Figure S2

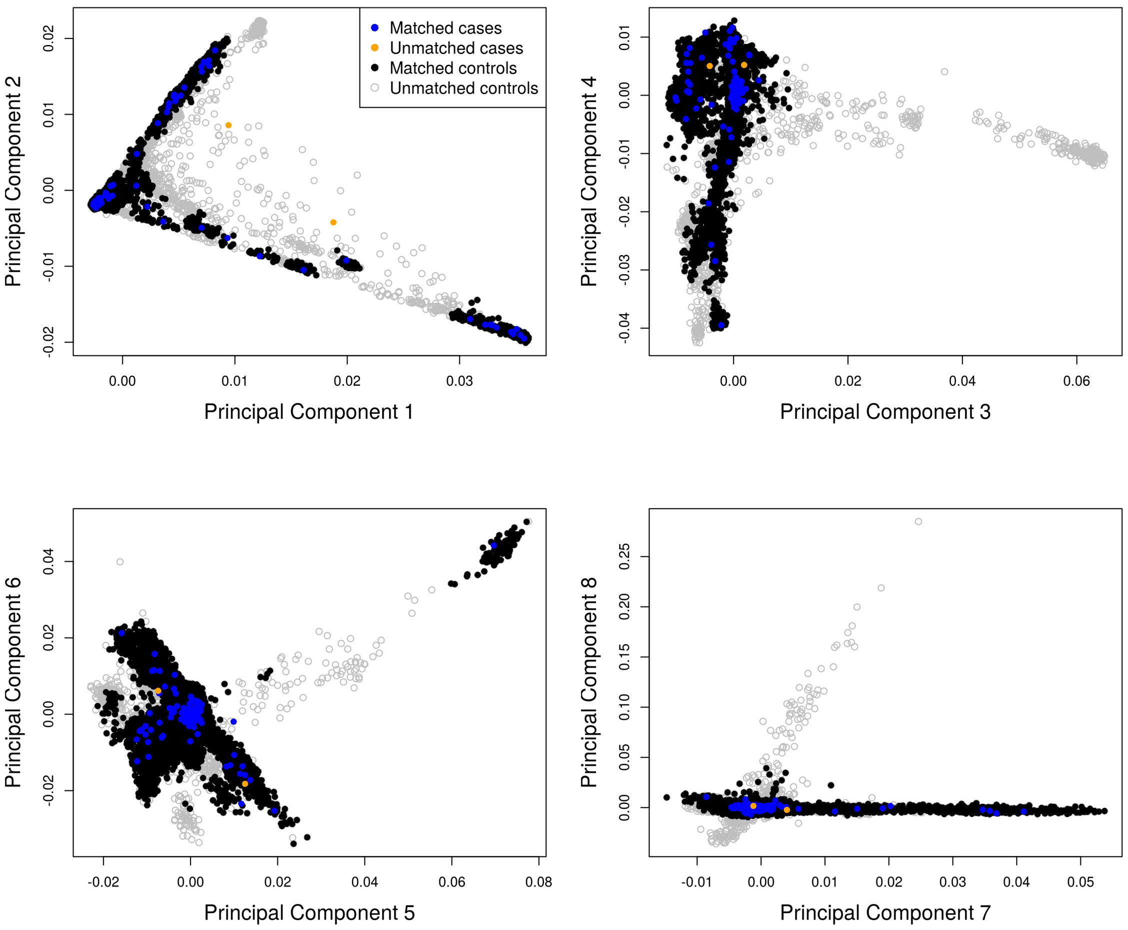


**Figure S2 | Ancestry matching.** Principal component analysis showing the first eight principal components for matched cases (blue) and controls (black) and unmatched cases (orange) and controls (grey). Two cases and 2,579 controls were excluded from downstream analyses.

#### Figure S3

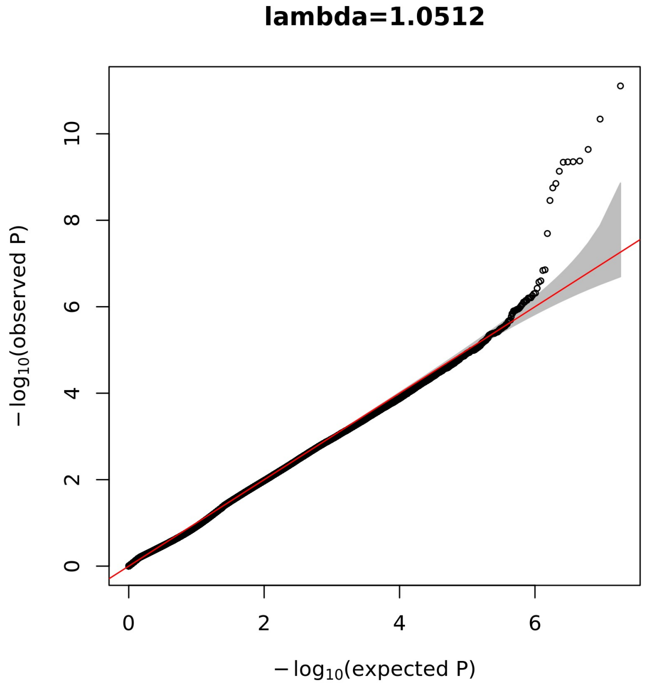


**Figure S3 | Q-Q plot for mixed-ancestry GWAS.** Quantile-quantile (Q-Q) plot displaying the observed versus the expected $-\log_{10}(P)$ for each variant tested. The grey shaded area represents the 95% confidence interval of the null distribution.

#### Figure S4

A

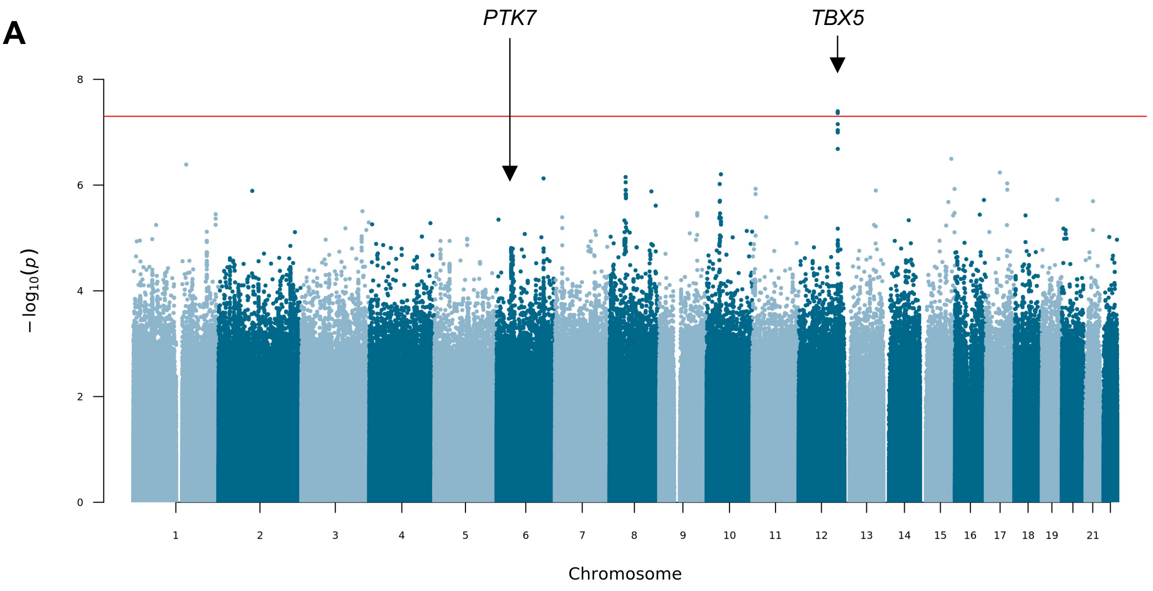


B

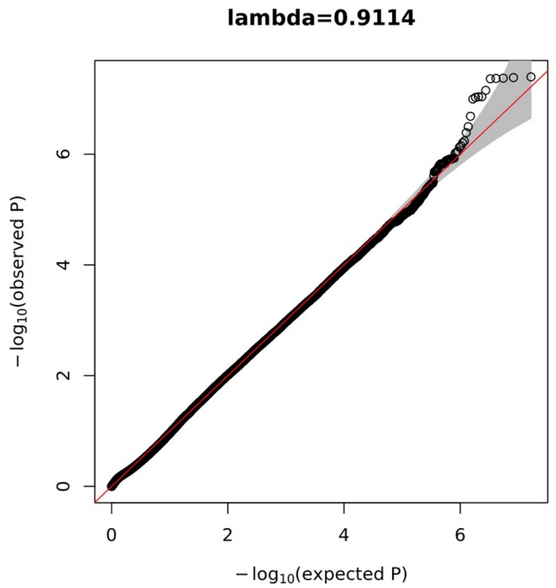


**Figure S4 | European GWAS.** A genome-wide single-variant association study was carried out in 88 cases and 17,993 controls for 15,447,192 variants with MAF > 0.1%. All cases and controls had genetically-determined European ancestry. **A**, Manhattan plot with chromosomal position (GRCh38) denoted along the *x* axis and strength of association using a $-\log_{10}(P)$ scale on the *y* axis. Each dot represents a variant. The red line indicates the Bonferroni adjusted threshold for genome-wide significance ($P < 5x10^{-8}$). The two genome-wide significant loci from the mixed ancestry GWAS are labelled. **B**, Quantile-Quantile (Q-Q) plot displaying the observed versus the expected $-\log_{10}(P)$ for each variant tested. The grey shaded area represents the 95% confidence interval of the null distribution.

#### Figure S5

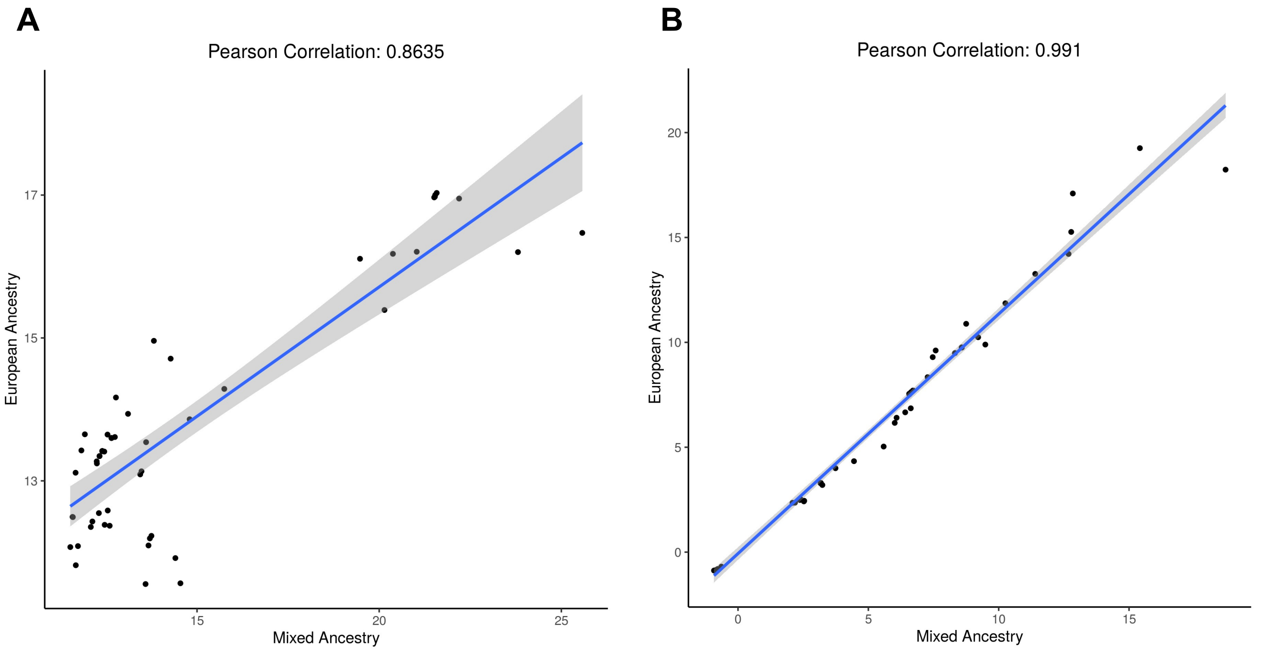


**Figure S5 | Correlation between mixed-ancestry and European GWAS.** Comparison of **A**, -$\log_{10}(P)$ and **B**, BETA from the mixed-ancestry and European-only ancestry GWAS. All variants with $P < 10^{-5}$ in both cohorts are shown. The shaded grey area represents the 95% confidence interval.

### Figure S6

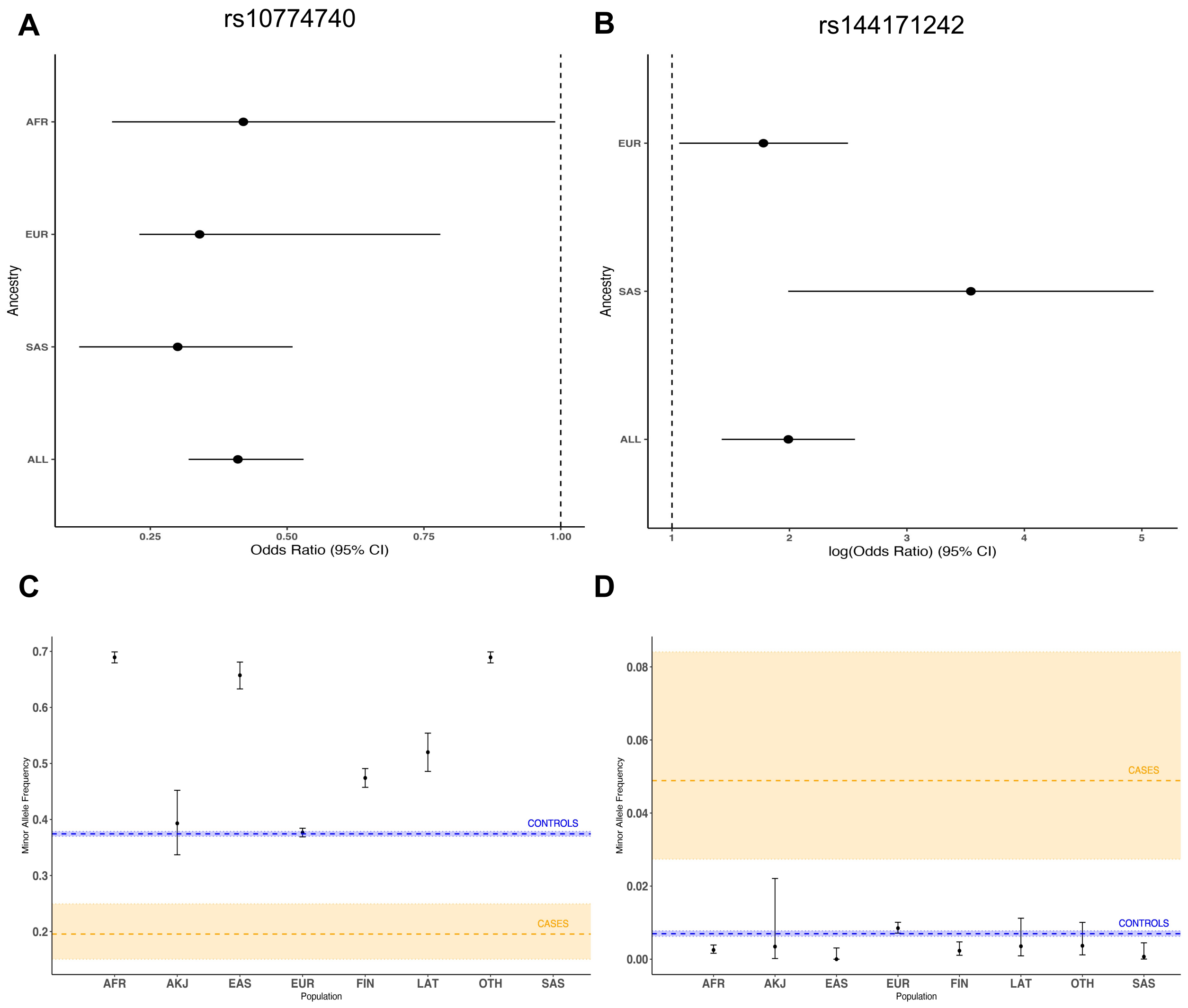


**Figure S6 | Comparison of ancestry-specific odds ratios and gnomAD allele frequencies.** GWAS per-ancestry odds ratios for **A**, rs10774740 and **B**, rs144171242. Comparison of population minor allele frequencies from gnomAD with case and control allele frequencies from our data for **C**, rs10774740 and **D**, rs144171242. Error bars represent 95% confidence intervals. The dashed lines indicate the minor allele frequency observed in cases (orange) and controls (blue) with the shaded areas indicating 95% confidence interval for each group. No data was available for rs10774740 in the South Asian population. AFR, African/African-American; AKJ, Ashkenazi Jewish; EAS, East Asian; EUR, European (non-Finnish); FIN, European (Finnish); LAT, Latino/Admixed-American; OTH, Other; SAS, South Asian.

#### Figure S7

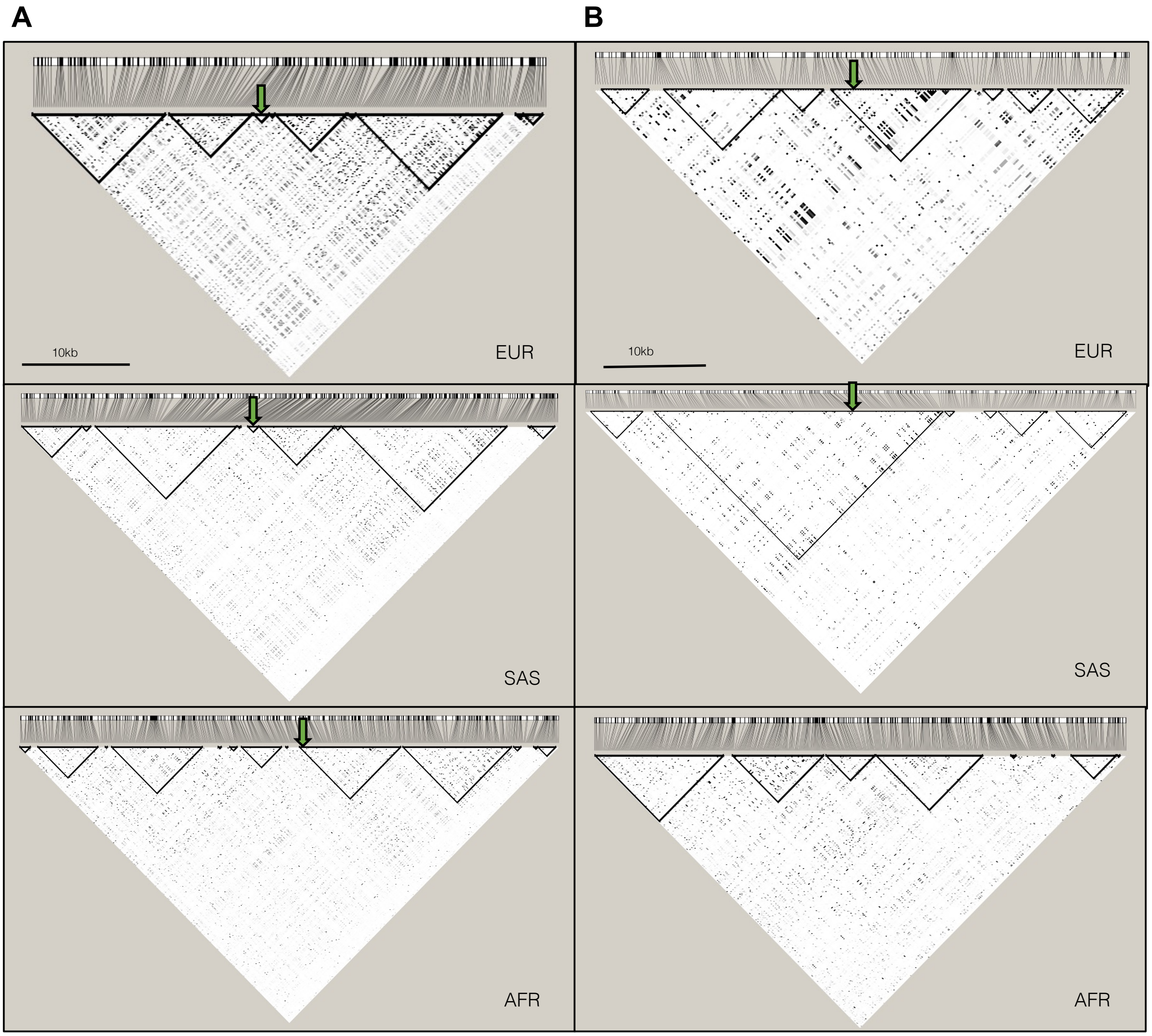


**Figure S7 | Linkage disequilibrium (LD) for reference populations in the 1000 Genomes Project.** LD plots for 503 European (EUR), 489 South Asian (SAS) and 661 African (AFR) ancestry individuals from the 1000 Genomes Project (Phase 3). Haploview (v4.2) was used to compute pairwise LD statistics ($r^2$) between variants for each population. The darker the shading, the higher the LD between variants. Black outlined triangles indicate haploblocks. **A**, LD plot for chr12:114,641,202-114,691,202 (GRCh37) with the position of the lead variant rs10774740 represented by a green arrow; **B**, LD plot for chr6:43,063,094-43,113,094 (GRCh37) with the position of the lead variant rs144171242 represented by a green arrow. rs144171242 was not seen in the AFR population group.

### Figure S8

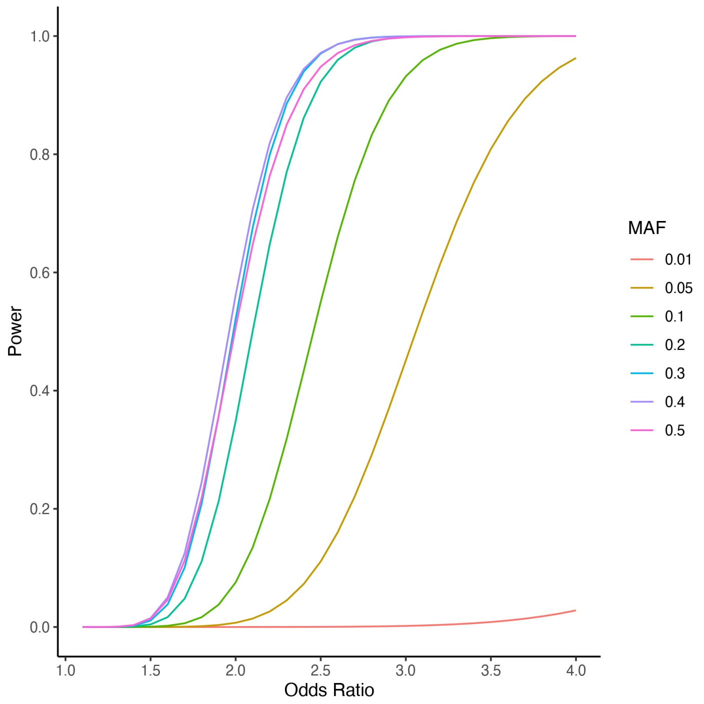


**Figure S8 | GWAS power calculation.** Power calculations were performed at various minor allele frequencies (MAF) using 132 cases and 23,727 controls under an additive genetic model to achieve genome-wide significance of $P<5x10^{-8}$.

**Table S1**. **Clinical characteristics and genetic ancestry**. PUV, posterior urethral valves; PCA, principal components analysis; EUR, European; SAS, South Asian; AFR, African; AMR, Latino/Admixed American; VUR, vesico-ureteral reflux; UTI, urinary tract infection; ESRD, end-stage renal disease.

| | | PUV (n=132) | Controls (n=23,727) |
|---|---|---|---|
| **Median age (range)** | | 13 (2-66) | |
| **Males (%)** | | 132 (100) | 10,425 (43.9) |
| **PCA determined ancestry** | | | |
| | EUR (%) | 89 (67.4) | 19,418 (81.8) |
| | SAS (%) | 18 (13.6) | 2847 (12.0) |
| | AFR (%) | 11 (8.3) | 449 (1.9) |
| | AMR (%) | 0 (0) | 7 (0.03) |
| | Admixed (%) | 14 (10.6) | 1006 (4.2) |
| **Additional renal/urinary phenotypes** | | | |
| | Hydronephrosis (%) | 56 (42.4) | |
| | Bladder abnormality (%) | 32 (24.2) | |
| | Hydroureter (%) | 30 (22.7) | |
| | VUR (%) | 27 (20.5) | |
| | Renal dysplasia (%) | 16 (12.1) | |
| | Hypertension (%) | 11 (8.3) | |
| | Renal agenesis (%) | 8 (6.1) | |
| | Recurrent UTIs (%) | 5 (3.8) | |
| | Renal hypoplasia (%) | 4 (3.0) | |
| | Renal duplication (%) | 2 (1.5) | |
| **Extrarenal manifestations (%)** | | 35 (26.5) | |
| | Cardiac anomaly (%) | 4 (3.0) | |
| | Neurodevelopmental disorder (%) | 7 (5.3) | |
| **Family history (%)** | | 5 (3.8) | |
| **End-stage renal disease (%)** | | 23 (17.4) | |
| **Median age ESRD (range)** | | 14 (0-39) | |

**Table S2**. **Exome-wide rare SNV/indel analysis**. Results from SAIGE-GENE aggregate rare (MAF < 0.1%) coding variant association. Gene name and Ensembl identifier listed for all genes with $P < 0.01$. See supplemental data.

**Table S3. Structural variant analysis.** The burden of rare autosomal structural variants intersecting with a) at least one exon or b) a cis-regulatory element was compared between 132 cases and 23,727 controls. cCRE, candidate cis-regulatory element; PUV, posterior urethral valves; CNV, copy number variant; DEL, deletion; DUP, duplication; INV, inversion; OR, odds ratio; CI, 95% confidence interval, IQR, interquartile range.

| | | EXON | | cCRE | |
|---|---|---|---|---|---|
| | | **PUV** | **Controls** | **PUV** | **Controls** |
| **CNV** | **n (%)** | 109 (82.6) | 17,961 (75.7) | 111 (84.1) | 18,773 (79.1) |
| | **OR (CI)** | 1.52 (0.96-2.50) | | 1.39 (0.87-2.34) | |
| | **Fisher's exact P** | 0.07 | | 0.20 | |
| | **Median size (kb) (IQR)** | 104 (183) | 94 (158) | 80 (165) | 80 (128) |
| | **P (Wilcoxon)** | 0.75 | | 0.75 | |
| **DEL** | **n (%)** | 117 (88.6) | 19,987 (84.2) | 132 (100) | 23,031 (97.1) |
| | **OR (CI)** | 1.46 (0.85-2.69) | | Inf | |
| | **Fisher's exact P** | 0.19 | | 0.04 | |
| | **Median size (kb) (IQR)** | 1.4 (4.2) | 1.8 (4.6) | 1.5 (4.1) | 1.9 (4.5) |
| | **P (Wilcoxon)** | 0.18 | | $4.1 \times 10^{-4}$ | |
| **DUP** | **n (%)** | 59 (44.7) | 8,476 (35.7) | 104 (78.8) | 16,004 (67.5) |
| | **OR (CI)** | 1.45 (1.01-2.08) | | 1.79 (1.17-2.83) | |
| | **Fisher's exact P** | 0.04 | | $5.0 \times 10^{-3}$ | |
| | **Median size (kb) (IQR)** | 3.7 (5.7) | 3.0 (5.6) | 2.0 (5.3) | 2.3 (5.3) |
| | **P (Wilcoxon)** | 0.16 | | 0.49 | |
| **INV** | **n (%)** | 66 (50.0) | 8,736 (36.8) | 81 (61.4) | 11,171 (47.1) |
| | **OR (CI)** | 1.72 (1.20-2.45) | | 1.79 (1.24-2.59) | |
| | **Fisher's exact P** | $2.1 \times 10^{-3}$ | | $1.2 \times 10^{-3}$ | |
| | **Median size (kb) (IQR)** | 253 (1931) | 261 (1642) | 129 (459) | 94 (779) |
| | **P (Wilcoxon)** | 0.44 | | 0.12 | |

**Table S4. Exome-wide gene-based structural variant analysis**. The burden of rare (AF < 0.1%), autosomal structural variants intersecting with exons was compared across 19,907 genes (GENCODE v29). Only genes intersecting with structural variants are listed. *P* values were derived using a two-sided Fisher's exact test. CNV, copy number variant; DEL, deletion; DUP, duplication; INV, inversion; SV, structural variant. See supplemental data.

**Table S5. Structural variant cCRE analysis.** The burden of rare autosomal structural variants intersecting with each cis-regulatory element type was compared between 132 cases and 23,727 controls. cCRE, candidate cis-regulatory element; CNV, copy number variant; DEL, deletion; DUP, duplication; INV, inversion; dELS, distal enhancer-like signature; pELS, proximal enhancer-like signature; PLS, promoter-like signature; OR, odds ratio; CI, 95% confidence interval.

| | | CNV | DEL | DUP | INV |
|---|---|---|---|---|---|
| **dELS** | No of cases (%) | 109 (82.6) | 130 (98.5) | 91 (68.9) | 75 (56.8) |
| | No of controls (%) | 18,546 (78.2) | 22,743 (95.9) | 13,727 (57.9) | 10,291 (43.4) |
| | Fisher's exact P | 0.25 | 0.18 | 0.01 | $2.0 \times 10^{-3}$ |
| | OR (CI) | 1.32 (0.84-2.18) | 2.81 (0.76-23.5) | 1.62 (1.11-2.40) | 1.72 (1.20-2.47) |
| **pELS** | No of cases (%) | 95 (72.0) | 88 (66.7) | 39 (29.5) | 57 (43.2) |
| | No of controls (%) | 14,755 (62.2) | 13,446 (56.7) | 3,988 (16.8) | 6,848 (28.9) |
| | Fisher's exact P | 0.02 | 0.02 | $2.7 \times 10^{-4}$ | $4.9 \times 10^{-4}$ |
| | OR (CI) | 1.56 (1.06-2.35) | 1.53 (1.05-2.25) | 2.08 (1.39-3.05) | 1.87 (1.30-2.68) |
| **PLS** | No of cases (%) | 82 (62.1) | 39 (29.5) | 11 (8.3) | 48 (36.4) |
| | No of controls (%) | 12,382 (52.2) | 6,243 (26.3) | 1,557 (6.6) | 5,862 (24.7) |
| | Fisher's exact P | 0.02 | 0.43 | 0.38 | $3.2 \times 10^{-3}$ |
| | OR (CI) | 1.50 (1.04-2.18) | 1.17 (0.79-1.73) | 1.29 (0.63-2.41) | 1.74 (1.19-2.52) |
| **CTCF-only** | No of cases (%) | 101 (76.5) | 79 (59.8) | 24 (18.2) | 65 (49.2) |
| | No of controls (%) | 16,247 (68.5) | 12,124 (51.1) | 3,368 (14.2) | 7,512 (31.7) |
| | Fisher's exact P | 0.05 | 0.05 | 0.21 | $3.1 \times 10^{-5}$ |
| | OR (CI) | 1.50 (0.99-2.32) | 1.43 (0.99-2.06) | 1.34 (0.82-2.11) | 2.09 (1.46-2.99) |
| **DNase-H3K4me3** | No of cases (%) | 86 (65.2) | 52 (39.3) | 19 (14.4) | 52 (39.4) |
| | No of controls (%) | 13,567 (57.2) | 7,298 (30.8) | 1,976 (8.3) | 6,397 (27.0) |
| | Fisher's exact P | 0.08 | 0.04 | 0.02 | $2.2 \times 10^{-3}$ |
| | OR (CI) | 1.40 (0.97-2.05) | 1.46 (1.01-2.10) | 1.85 (1.07-3.03) | 1.76 (1.22-2.53) |

**Table S6. Mixed ancestry GWAS association statistics.** Summary statistics for all variants with $P < 10^{-5}$. Allele 2 refers to the effect allele. Allele 1 is the other allele. CHR, chromosome; POS, genomic position with reference to GRCh38; SE, standard error; AF, allele frequency. See supplemental data.

**Table S7. GWAS gene-based analysis.** MAGMA was used to assess the joint effect of common and low-frequency variants across genes. Genes with $P < 0.01$ are listed. CHR, chromosome; START and STOP denote the genomic position with reference to GRCh37; NSNPS; the number of variants aggregated for each gene. See supplemental data.

**Table S8. GWAS pathway analysis.** MAGMA was used to assess the joint effect of common and low-frequency variants across different biological pathways. Pathways with $P < 0.05$ are listed. NGENES; the number of genes aggregated across each pathway; SE, standard error; GO, gene ontology. See supplemental data.

**Table S9. Replication study.** The lead variants at the top four loci with $P < 5x10^{-7}$ were genotyped in an independent European cohort of 398 PUV cases and 10,804 controls. *P* values in the replication cohort were calculated using a one-sided Cochran Armitage Trend test. OR, odds ratio; CI, 95% confidence interval.

| Locus | Lead variant | Effect Allele | Discovery *P* value | Replication *P* value | Discovery OR (95% CI) | Replication OR (95% CI) |
|---|---|---|---|---|---|---|
| 12q24.21 | rs10774740 | T | $7.81x10^{-12}$ | $1.9x10^{-3}$ | 0.40 (0.31-0.52) | 0.78 (0.67-0.91) |
| 6p.21.1 | rs144171242 | G | $2.02x10^{-8}$ | $4.5x10^{-3}$ | 7.20 (4.08-12.70) | 2.17 (1.25-3.76) |
| 10q11.21 | rs1471950716 | A | $1.45x10^{-7}$ | NA | 3.88 (2.42-6.22) | NA |
| | rs137855548 | G | $1.46x10^{-6}$ | 0.5471 | 3.94 (2.36-6.56) | 0.84 (0.48-1.47) |
| 14q21.1 | rs199975325 | G | $2.52x10^{-7}$ | 0.9636 | 5.68 (3.22-9.99) | 1.02 (0.52-1.98) |

**Table S10. Comparison of mixed ancestry and European GWAS association statistics.** The lead variants at the top four loci with $P < 5x10^{-7}$ are shown. OR, odds ratio; CI, 95% confidence interval.

| Lead variant | Effect Allele | Mixed ancestry *P* value | European *P* value | Mixed ancestry OR (95% CI) | European OR (95% CI) |
|---|---|---|---|---|---|
| rs10774740 | T | $7.81x10^{-12}$ | $7.03x10^{-8}$ | 0.40 (0.31-0.52) | 0.42 (0.10-0.73) |
| rs144171242 | G | $2.02x10^{-8}$ | $3.60x10^{-5}$ | 7.20 (4.08-12.70) | 5.90 (2.88-12.11) |
| rs1471950716 | A | $1.45x10^{-7}$ | $6.24x10^{-5}$ | 3.88 (2.42-6.22) | 4.44 (2.68-7.36) |
| rs199975325 | G | $2.52x10^{-7}$ | $1.57x10^{-5}$ | 5.68 (3.22-9.99) | 4.43 (2.16-9.06) |
